# Aldosterone Suppression Testing and Subtype Prediction for Primary Aldosteronism

**DOI:** 10.64898/2026.05.14.26353176

**Authors:** Manaporn Payanundana, Wasita W. Parksook, Kantawich Piyanirun, Dutsadee Charunvarakornchai, Chonpiti Siriwan, Stefanie Parisien-La Salle, Cheng-Hsuan Tsai, Andrew J. Newman, Jenifer M. Brown, Nattapol Sathavarodom, Sarat Sunthornyothin, Apussanee Boonyavarakul, Anand Vaidya

**Affiliations:** Division of Endocrinology and Metabolism, Department of Medicine, Phramongkutklao Hospital, and Phramongkutklao College of Medicine, Bangkok, Thailand; Center for Adrenal Disorders, Division of Endocrinology, Diabetes, and Metabolism, Brigham and Women’s Hospital, Harvard Medical School, Boston, MA, USA; Division of Endocrinology and Metabolism, Thai Red Cross Society, Bangkok, Thailand; Department of Medicine, Faculty of Medicine, Chulalongkorn University, Thai Red Cross Society, Bangkok, Thailand; Excellence Center for Diabetes, Hormones and Metabolism, King Chulalongkorn Memorial Hospital, Thai Red Cross Society, Bangkok, Thailand; Division of General Internal Medicine, Department of Medicine, Faculty of Medicine, Chulalongkorn University and King Chulalongkorn Memorial Hospital, Thai Red Cross Society, Bangkok, Thailand; Division of Endocrinology, Department of Medicine, Centre Hospitalier de l’Université de Montréal, Université de Montréal, QC, Canada; Division of Cardiology, Department of Internal Medicine, National Taiwan University Hospital and National Taiwan University College of Medicine, Taipei 100, Taiwan; Primary Aldosteronism Center at National Taiwan University Hospital, Taipei 100, Taiwan; Heart & Vascular Institute, Brigham and Women’s Hospital, Harvard Medical School, Boston, MA, USA

## Abstract

**Objectives:** To evaluate the accuracy of predicting PA subtype using: 1) the 2025 Endocrine Society primary aldosteronism (PA) guideline-endorsed classification to “high,” “intermediate,” and “low” probabilities of lateralization; and 2) the seated saline suppression test (SST) results.

**Design:** A multicenter, retrospective, cohort study

**Methods:** The discriminatory capacity of guideline-endorsed probability frameworks for PA subtyping were evaluated in this cohort of 319 PA patients, from two large tertiary centers in Bangkok, Thailand, who underwent subtyping assessments regardless of probability status. PA subtypes were determined by adrenal venous sampling (AVS) and/or post-adrenalectomy outcomes using PASO criteria.

**Results:** The majority of PA patients were characterized as having “intermediate” probability for lateralizing PA (75%); however, lateralizing PA was ultimately confirmed in 61-78% of all patients, regardless of guideline-based probability classification. The vast majority of SST results were positive using guideline-derived criteria, regardless of probability stratification or ultimate subtype: 89.3% of patients with lateralizing PA and 80.6% of those with bilateral PA had a positive SST. Among patients with “intermediate” probability of lateralizing PA, where guidelines specifically endorse the value of SST, the SST had a sensitivity of 89.4% and specificity of 22.0% for detecting lateralizing PA, with 78.0% false-positive and 10.6% false-negative rates. Consistently, post-SST aldosterone concentrations exhibited near-complete overlap between those with and without lateralizing PA.

**Conclusion:** Guideline-endorsed probability frameworks, and the use of SST, lacked discriminatory capacity to predict PA subtype.

## Introduction

Primary aldosteronism (PA) is characterized by renin-independent aldosterone production that manifests across a phenotypic severity spectrum^1–3^. PA is a common cause of hypertension and cardiorenal disease^4–7^ that can be mitigated with disease-specific medical or surgical interventions^8^. The current management of PA depends on the subtype: a minority of patients have lateralizing PA that would benefit from unilateral adrenalectomy, whereas the vast majority of patients have non-lateralizing PA for which medical therapy is indicated and can be very effective^4,9,10^.

In clinical practice, adrenal venous sampling (AVS) remains the most widely accepted method for determining PA subtype^4,11,12^. However, its limited availability, high cost, and resource-intensive nature render it a bottleneck in PA evaluation. To address this bottleneck, the 2025 Endocrine Society (ES) created new approaches to stratify the probability of lateralization for each patient using biochemical values (for renin, aldosterone, and potassium). In this updated approach, the ES guidelines recommended treatment approaches based on whether patients with PA have a “low”, “intermediate”, or “high” probability of lateralization. This guideline suggests that patients classified as having a “low” probability for lateralizing PA be treated medically without further testing, whereas those deemed to have a “high” probability be referred for AVS. Further, a new guideline suggestion that emerged was to pursue any one of three aldosterone suppression tests when the probability classification was “intermediate,” with the assumption that the results would help triage the appropriate use of AVS: a positive test result is assumed to identify those who are likely to lateralize on AVS and thus justify the pursuit of this resource, whereas a negative test is assumed to identify those unlikely to lateralize on AVS and for whom medical therapy should instead be pursued. Because this new guideline approach was based on expert consensus derived from limited empirical data or rigorous clinical validation^13^, and because the performance of aldosterone suppression tests to discriminate PA subtype has recently been shown to be limited across multiple cohorts^14–18^, we sought to examine the reliability and accuracy of these guideline recommendations in a large multi-site cohort of Asian PA patients.

Herein, we evaluated the discriminatory performance of the ES guideline-based probability stratifications for predicting lateralizing PA (low, intermediate, high) and the performance of aldosterone suppression testing (using the seated saline suppression test [SST]) for determining lateralizing PA in a large cohort of Thai patients with PA who underwent subtyping procedures regardless of the results of their probability stratification or aldosterone suppression testing.

## Methods

### Study Design and Population

This was a multicenter, retrospective, cohort study including patients diagnosed with PA at two large tertiary referral centers in Bangkok, Thailand. Consecutive patients with PA were included from Phramongkutklao Hospital (2016-2025) and King Chulalongkorn Memorial Hospital (2020-2025). The objective of this study was to assess the diagnostic performance of the newly proposed criteria to determine the probability of lateralizing PA in the 2025 ES guidelines. The 2025 ES guidelines recommend ascertainment of the probability of the ultimate PA subtype by categorizing patients as “high”, “intermediate”, or “low” probability for lateralizing PA using biochemical criteria, and subsequently, recommend using aldosterone suppression testing for those in the “intermediate” probability subgroup to ascertain eligibility for AVS^4^.

Eligible patients were at least 18 years of age and had a diagnosis of PA established between 2016–2025 using contemporary guidelines^19^. All patients had positive screening tests and underwent aldosterone suppression testing unless they exhibited spontaneous hypokalemia with undetectable renin and plasma aldosterone concentration (PAC) ≥550 pmol/L (20 ng/dL). Patients with post-SST PAC >6 ng/dL were considered as having a positive result consistent with PA^19^; patients with a post-SST PAC ≤6 ng/dL were classified as having PA or not based on expert clinical judgment^14–16,18^. Frequently, despite a post-SST PAC ≤6 ng/dL implying a “negative” result, clinicians still made a diagnosis of PA and overruled the aldosterone suppression test due to severe renin-independent aldosteronism, and/or hypokalemia, and/or the severity of the blood pressure phenotype.

For the purpose of this retrospective re-interpretation analysis, these historically confirmed cases were then stratified post hoc into the "high", "intermediate", or "low" probability subgroups as defined by the newly proposed 2025 ES criteria. Additionally, all eligible individuals must have undergone adrenal imaging with computed tomography (CT) or magnetic resonance imaging (MRI), and had definitive ascertainment of their PA subtype by either AVS and/or surgical and biochemical follow-up after unilateral adrenalectomy using the standardized Primary Aldosteronism Surgical Outcome (PASO) criteria^20^ (**Figure 1**). Two complementary analyses were performed to evaluate the performance and applicability of the 2025 ES guideline recommendations for predicting PA subtype:

1. Prediction of the probability of lateralizing PA as “low”, “intermediate”, or “high”, using renin, aldosterone, and potassium values as proposed by the 2025 ES guidelines. All 319 patients were eligible for this analysis.
2. Diagnostic performance of the seated SST for predicting lateralizing PA, to assess the accuracy of relying on aldosterone suppression testing for this purpose, as proposed by the 2025 ES guidelines. From the cohort of 319 patients, 188 (59%) had undergone an SST with subsequent subtyping procedures, irrespective of the SST result, and were thus eligible for this analysis.

**Figure 1.**
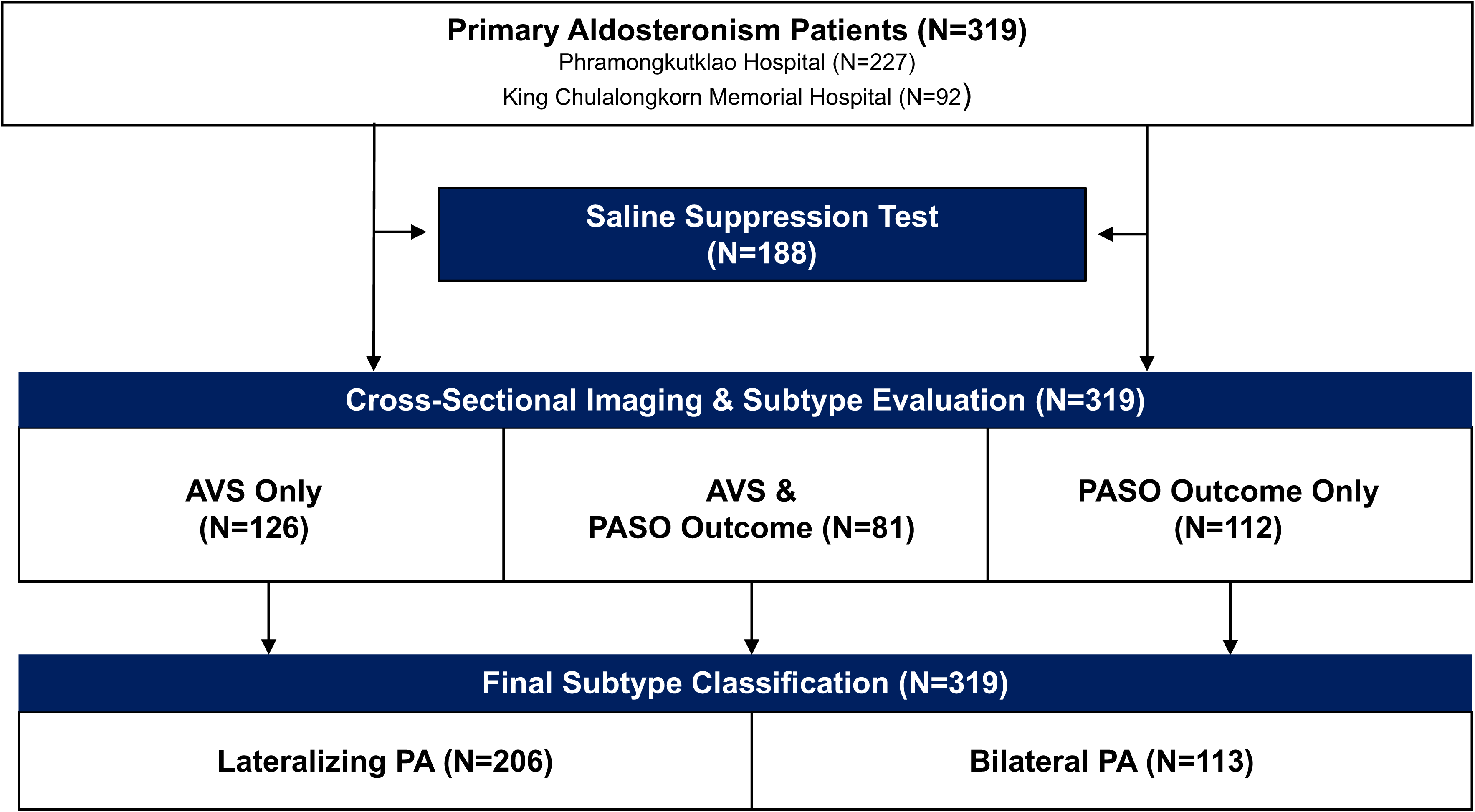
Flowchart of the Study Population and Methods for Subtype Evaluation. Abbreviations: AVS, adrenal venous sampling; PASO, primary aldosteronism surgical outcome.

A key feature of this study is that it leveraged the fact that practicing clinicians routinely followed prevailing ES guidelines for the evaluation of PA, but in addition, routinely pursued subtyping procedures and clinical treatment decisions that overruled or bypassed traditional guidance. As a result, this study was able to include PA patients who had subtype ascertainment regardless of whether their probability classification was “low,” “intermediate”, or “high,” and regardless of their SST results, including when SST was “negative”. As such, this study was able to assess 2×2 accuracy tables (including positive and negative outcomes) that permitted assessment of the discriminatory capacity of guideline-endorsed subtype prediction approaches.

The study was approved by the Institutional Review Board of both hospitals and conducted in accordance with the Declaration of Helsinki. A waiver of informed consent was granted by the Institutional Review Board.

### Clinical Evaluation and Management of Interfering Medications

Prior to conducting screening tests for PA, interfering antihypertensive medications were discontinued for at least 2-4 weeks, when feasible, and patients remained on the same medications until the completion of the SST. Patients were treated with alpha blockers, hydralazine, and/or non-dihydropyridine calcium channel blockers for blood pressure control during this time.

### Assessment of Probability of Lateralizing PA

Patients were classified according to the 2025 ES probability criteria as follows, strictly adhering to the detailed criteria outlined in the Technical Remarks of the 2025 guideline:

- “Low” probability of lateralizing PA: Normokalemia (serum potassium ≥ 3.5 mmol/L) and plasma aldosterone concentration (PAC) ≤ 305 pmol/L (11 ng/dL) by immunoassay
- “Intermediate” probability of lateralizing PA comprised all patients who did not fulfill the criteria for either “high” or “low” probability of lateralization.
- “High” probability of lateralizing PA: Resistant hypertension or hypertension with PAC > 554 pmol/L (20 ng/dL) by immunoassay and PRA < 0.2 ng/mL/h, and the presence of hypokalemia.

### Seated Saline Suppression Test (SST)

All SSTs were performed between 08:00 and 10:00 AM. Serum potassium was corrected with supplementation prior to performing the SST to ensure normokalemia for the procedure. Patients maintained a seated position for 30 minutes prior to the infusion of two liters of normal saline over four hours, followed by the measurement of PAC at the end of the infusion^4^. As per the 2025 ES guidelines^4^, the seated SST was considered “positive” if post-infusion PAC ≥ 217 pmol/L (7.8 ng/dL), and “negative” if post-infusion PAC was below this threshold.

### Subtype Determinations

PA subtype was determined by one of two methods:

1. AVS using the criteria described below^11,12^; and/or
2. Biochemical outcomes following unilateral adrenalectomy (**Table 1).** For patients the minority of patients who underwent unilateral adrenalectomy prior to 2018 (n=53) (before the publication of the PASO criteria^20^), post-operative outcomes were determined clinically, including resolution or significant improvement of hypertension and hypokalemia without biochemical measures. Additional sensitivity analyses, described below, were conducted after excluding individuals prior to 2018 to ensure analytic rigor.

**Table 1.** Definitions of Clinical and Biochemical Outcomes based on the PASO Criteria.

| Outcome assessment | Clinical outcomes | Biochemical outcomes |
| --- | --- | --- |
| **Complete success** | Normal blood pressure (BP < 140/90 mmHg) without using the antihypertensive medication | Normalization of potassium and normalization of the PAC < 138.7 pmol/L (5 ng/dL) by immunoassay, or aldosterone-to-renin ratio (if available). |
| **Partial success** | The same blood pressure as before surgery with less antihypertensive medication or a reduction in blood pressure with either the same amount or less antihypertensive medication | Normalized potassium with reduced and a raised aldosterone-to-renin ratio (if available) with one or both of the following: ≥50% decrease in baseline PAC; or abnormal but improved post-surgery aldosterone suppression test. |

Abbreviations: BP, Blood pressure; PAC; Plasma aldosterone concentration.

Lateralizing PA was diagnosed based on either 1) having lateralization on AVS (as defined below) and/or 2) achieving biochemical improvement following unilateral adrenalectomy (Supplemental Table 1), including in some patients who went directly to surgery without AVS. Those with post-operative PAC ≥10 ng/dL with suppressed renin following unilateral adrenalectomy were considered as having bilateral PA. In a small minority of cases where there were discordant results between AVS and post-operative biochemical outcomes, classification was based on post-operative biochemical outcomes (**Table 2**).

**Table 2.** Individuals with Discordant Results Between AVS and PASO Outcomes.

| Subtype | Patient<br>no. | Probability<br>Group | AVS<br>Protocol | AVS Results | Biochemical<br>Success |
| --- | --- | --- | --- | --- | --- |
| **Lateralizing (n= 7)** | 1 | Intermediate | Post | No lateralization | Complete |
|  | 2 | Intermediate | Post | No lateralization | Complete |
|  | 3 | Intermediate | Post | No lateralization | Complete |
|  | 4 | High | Post | No lateralization | Complete |
|  | 5 | High | Post | No lateralization | Complete |
|  | 6 | High | Post | No lateralization | Complete |
|  | 7 | High | Pre | Bilateral suppression | Complete |
| **Bilateral PA (n= 9)** | 1 | Low | Pre | Lateralization | Absence |
|  | 2 | Intermediate | Post | Lateralization | Absence |
|  | 3 | Intermediate | Pre | Lateralization | Absence |
|  | 4 | Intermediate | Pre | Lateralization | Absence |
|  | 5 | Intermediate | Pre | Lateralization | Absence |
|  | 6 | Intermediate | - | - | Absence |
|  | 7 | High | Pre | Lateralization | Absence |
|  | 8 | High | - | - | Absence |
|  | 9 | High | Post | Lateralization | Absence |

AVS Protocol indicates the timing of adrenal venous sampling relative to cosyntropin administration. Pre: Pre-ACTH stimulation (baseline sampling). Post: Post-ACTH stimulation (sampling during or after continuous cosyntropin infusion). - (Dash): Data not available or not applicable.

### Adrenal Venous Sampling (AVS)

AVS was performed according to each institution’s local protocol during the respective study periods as outlined below. Although AVS capability was available at all participating institutions, it could not be performed in all patients due to patient-related factors, willingness for surgery, and resource availability constraints^12,21,22^.

At Phramongkutklao Hospital (from 2016-2025) and at King Chulalongkorn Memorial Hospital (from 2020-2021), AVS was performed with continuous adrenocorticotropic hormone (ACTH) stimulation. ACTH was infused into a peripheral vein at a rate of 50 mcg/h, starting 30 minutes before sampling. Successful cannulation was defined as a selectivity index ≥ 5 after stimulation. Lateralizing on AVS was defined as a lateralization index ≥ 4 after stimulation.

After 2021, AVS at King Chulalongkorn Memorial Hospital was performed using a sequential technique without cosyntropin stimulation. Successful cannulation was defined as a selectivity index ≥ 2 without stimulation. Lateralization on AVS was defined as a lateralization index ≥ 2 without ACTH stimulation.

### Unilateral Adrenalectomy and PASO Outcomes

Among the 206 patients who were diagnosed with lateralizing PA, 171 underwent unilateral adrenalectomy. Blood pressure, serum potassium, PAC, and serum creatinine were recorded at baseline and postoperatively. Biochemical and clinical outcomes were evaluated according to PASO criteria within 1-year post-adrenalectomy^20^.

Among the 113 patients who were considered to have bilateral PA, 22 underwent unilateral adrenalectomy. The clinical pathways for these 22 patients were as follows: 8 patients had initial AVS results suggesting lateralization; 3 patients with bilateral AVS results still proceeded to unilateral adrenalectomy due to severe clinical indications^23^; 8 patients underwent unilateral adrenalectomy without AVS guided by clinical parameters and cross-sectional imaging findings; and 3 patients proceeded to unilateral adrenalectomy following failed AVS attempts. In each of these 19 cases, postoperative biochemical classifications confirmed improved but persistent PA. Postoperative outcomes to assess PASO criteria were available for 193 patients (**Figure 1**).

### Laboratory Measurements

The same clinical assays were consistently used for all reported measurements at both institutions. Aldosterone and renin testing was performed within the same laboratory, and immunoassay-based diagnostic thresholds were applied in the reported analyses. PAC was measured using a competitive radioimmunoassay (DIAsource ImmunoAssays S.A.; lowest reportable value was 69.3 pmol/L (2.5 ng/dL); interassay coefficient of variation [CV] = 6.2-18.5%; intra-assay CV = 5.3-13.7%; RRID: AB_2916289). PRA was measured by radioimmunoassay (DIAsource ImmonoAssays S.A.; interassay CV = 5.9-9.5%; intra-assay CV = 4.5-6.5%) (RRID: AB_2736926).

## Statistical Analysis

Continuous variables are presented as mean and standard deviation (SD) for normally distributed data and median (interquartile range, IQR) for non-normally distributed data. Diagnostic performance was evaluated using 2×2 contingency tables, from which sensitivity, specificity, positive predictive value (PPV), and negative predictive value (NPV) were calculated. Group comparisons used ANOVA, Kruskal-Wallis, or Chi-square tests, as appropriate. Fisher’s exact test was used to compare unilateral and bilateral PA across probability groups. Pre- and post-treatment changes were analyzed using paired *t*-tests or Wilcoxon signed-rank tests.

Beyond standard diagnostic metrics, the concordance between SST results and determined subtypes was analyzed both categorically, which classified SST results as positive or negative based on established cutoffs in 2×2 contingency tables, and continuously, post-SST PAC values were visualized across subtypes.

Multiple sensitivity analyses were conducted to ensure the robustness of the results, including:

- Separate analyses in only those patients who had AVS-confirmed subtype determination and in only those who had subtype that was PASO-confirmed (biochemical outcomes).
- Separate analyses were conducted after excluding the minority of patients who had subtype determinations before 2018, prior to the publication of PASO criteria, to ensure the rigor of subtype adjudication.
- Analyses to evaluate the diagnostic performance of individual AVS protocols by evaluating results in those with ACTH-stimulated and non-stimulated AVS separately to determine whether the difference in protocol influenced subtype interpretation.
- Separate analyses were conducted in only those with serum potassium ≥ 4.0 mmol/L at the beginning of SST, to more robustly demonstrate that the results were not influenced by serum potassium concentrations.
- Because the definition of “intermediate” risk for lateralizing PA was defined by the exclusion of both the low- and high-probability criteria in the ES guidelines, separate analyses were conducted in the following subgroups to evaluate a variety of interpretations of what constitutes intermediate risk:

- The subgroup of patients with PAC 305-554 pmol/L (11-20 ng/dL), concurrent normokalemia (serum potassium ≥ 3.5 mmol/L), and no history of hypokalemia.
- The subgroup of patients with PAC 305-554 pmol/L (11-20 ng/dL) regardless of their potassium status.
- The subgroup of patients with normokalemia (serum potassium ≥ 3.5 mmol/L), and no history of hypokalemia.

A two-sided p-value <0.05 was considered statistically significant. All analyses were performed using R software (version 4.5.1; The R Foundation for Statistical Computing, 2025).

## Results

Baseline characteristics of the study population are shown in **Table 3**. A total of 319 PA patients had subtyping results (**Figure 1**), of whom 188 underwent SST. Subtype classification was determined by only AVS (n = 126), only PASO criteria (n = 112), or both AVS and PASO data (n = 81). Therefore, a total of 207 patients had subtype determined via AVS (with 105 determined to be lateralizing and 102 determined to have bilateral PA), and 193 patients had subtype confirmation via PASO criteria (with 171 confirmed to have lateralizing PA and 22 confirmed to have bilateral PA, based on their strict lack of post-adrenalectomy biochemical success (**Table 1**), and had a post-operative PAC ≥10 ng/dL paired with suppressed renin or persistent hypokalemia). In total, 206/319 patients (65%) were determined to have lateralizing PA and 113/319 (35%) were determined to have bilateral PA (**Figure 1**). These subtype determinations served as the reference standard for the reported analyses.

**Table 3.** Baseline Characteristics by Probability Group.

| Variable | “Low”<br>Probability<br>(n = 6) | “Intermediate”<br>Probability<br>(n = 241) | “High”<br>Probability<br>(n = 72) | *p*-value |
| --- | --- | --- | --- | --- |
| Age (years) | 55.3 ± 9.7 | 51.1 ± 12.9 | 51.5 ± 12.3 | 0.48 |
| Female (%) | 66.7 | 53.1 | 65.3 | 0.19 |
| K (mmol/L) | 4.12 ± 0.36 | 3.30 ± 0.61 | 2.81 ± 0.46 | <0.001 |
| PAC (ng/dL) | 8.3 (7.1-9.1) | 26.1 (17.4-37.0) | 34.0 (27.5-52.8) | <0.001 |
| PAC (pmol/L) | 230.4 (196.9-251.8) | 724.0 (482.7-1026.4) | 941.8 (763.5-1463.3) | <0.001 |
| PRA (mcg/L/h) | 0.30 (0.10-0.63) | 0.39 (0.22-0.63) | 0.10 (0.05-0.12) | <0.001 |
| ARR (ng/dL per mcg/L/h) | 38.7 (12.7-73.0) | 71.2 (38.6-137.0) | 456.0 (258.6-905.7) | <0.001 |
| ARR (pmol/L per ng/mcg/L/h) | 1149.6 (354.5-2061.8) | 1973.8 (1070.7-3687.1) | 12649.4 (7189.6-25123.2) | <0.001 |
| eGFR (mL/min/1.73m<sup>2</sup>) | 89.5 ± 19.8 | 85.2 ± 22.4 | 89.2 ± 21.6 | 0.73 |
| SBP (mmHg) | 135 ± 18 | 152 ± 18 | 155 ± 16 | 0.08 |
| DBP (mmHg) | 83 ± 9 | 91 ± 13 | 93 ± 11 | 0.74 |
| No. of Antihypertensive drugs | 2.2 ± 0.8 | 2.2 ± 1.0 | 2.2 ± 1.0 | 0.80 |

Data are presented as mean ± SD, median (IQR), or percentage. P-values were derived from ANOVA or the chi-square test. Potassium (K) levels represent the nadir values obtained prior to the initiation of potassium supplementation. Abbreviations: K, potassium; SBP, systolic blood pressure; DBP, diastolic blood pressure; BMI, body mass index; eGFR, estimated glomerular filtration rate; PAC, plasma aldosterone concentration; PRA, plasma renin activity; ARR, aldosterone-to-renin ratio.

### Treatment outcomes

A total of 301 out of 319 patients had available post-treatment outcome data. Both lateralizing PA treated with unilateral adrenalectomy and bilateral PA treated with MRA or unilateral adrenalectomy showed significant reductions in SBP and DBP and normalization of serum potassium (**Table 4**). Lateralizing PA patients who were treated with MRA achieved potassium normalization and a significant reduction in SBP. Among 121 patients with lateralizing PA who underwent unilateral adrenalectomy and had available postoperative PAC data, PAC levels decreased significantly from 837.7 (629.7-1109.6) to 92.1 (58.3-165.6) pmol/L [30.2 (22.7-40.0) ng/dL to 3.3 (2.1-6.0) ng/dL] (*p* < 0.001). Among the 22 patients with persistent PA post-adrenalectomy who had been classified as having bilateral PA, the median post-operative PAC remained elevated at 453.3 pmol/L [16.3 ng/dL].

**Table 4.** Treatment Outcomes according to PA Subtypes.

| Parameters | Treatment | n | Pre-treatment | Post-treatment | Difference* | p-value |
| --- | --- | --- | --- | --- | --- | --- |
| **SBP (mmHg)** |  |  |  |  |  |  |
| **Lateralizing PA** | MRA | 7 | 149 ± 8 | 135 ± 15 | -14 | 0.02 |
| **Lateralizing PA** | Surgery | 187 | 154 ± 18 | 130 ± 13 | -24 | <0.001 |
| **Bilateral PA** | MRA | 85 | 149 ± 15 | 133 ± 13 | -17 | <0.001 |
| **Bilateral PA** | Surgery | 21 | 146 ± 17 | 135 ± 16 | -11 | 0.02 |
| **DBP (mmHg)** |  |  |  |  |  |  |
| **Lateralizing PA** | MRA | 7 | 95 ± 6 | 86 ± 11 | -9 | 0.08 |
| **Lateralizing PA** | Surgery | 187 | 92 ± 12 | 81 ± 11 | -11 | <0.001 |
| **Bilateral PA** | MRA | 85 | 89 ± 12 | 83 ± 11 | -6 | <0.001 |
| **Bilateral PA** | Surgery | 21 | 90 ± 10 | 81 ± 11 | -9 | 0.01 |
| **Potassium (mmol/L)** |  |  |  |  |  |  |
| **Lateralizing PA** | MRA | 7 | 2.8 ± 0.3 | 4.1 ± 0.5 | 1.3 | 0.004 |
| **Lateralizing PA** | Surgery | 184 | 3.1 ± 0.6 | 4.2 ± 0.4 | 1.1 | <0.001 |
| **Bilateral PA** | MRA | 84 | 3.4 ± 0.5 | 4.2 ± 0.4 | 0.8 | <0.001 |
| **Bilateral PA** | Surgery | 22 | 3.3 ± 0.8 | 4.2 ± 0.3 | 0.9 | <0.001 |
| **eGFR (mL/min/1.73m<sup>2</sup>)** |  |  |  |  |  |  |
| **Lateralizing PA** | MRA | 7 | 93.9 ± 23.5 | 81.5 ± 18.8 | -11.7 | 0.024 |
| **Lateralizing PA** | Surgery | 182 | 87.6 ± 22.3 | 80.1 ± 22.3 | -7.5 | <0.001 |
| **Bilateral PA** | MRA | 84 | 84.2 ± 22.5 | 88.9 ± 71.1 | 4.7 | 0.554 |
| **Bilateral PA** | Surgery | 20 | 76.4 ± 17.7 | 74.4 ± 23.9 | -2.0 | 0.612 |

Data are presented as mean ± SD.
\*The mean difference represents the change from baseline to post-treatment follow-up.
p-values denote the comparison between pre-treatment and post-treatment measures using the paired t-test or Wilcoxon signed-rank test. Abbreviations: PA, Primary aldosteronism; MRA, Mineralocorticoid receptor antagonists; SBP, systolic blood pressure; DBP, diastolic blood pressure; eGFR, Estimated glomerular filtration rate

To assess diagnostic heterogeneity, outcomes were compared between patients diagnosed by successful AVS (n=70) and those diagnosed by clinical/biochemical follow-up (n=51). There was no significant difference in post-operative PAC levels between the two groups (Median 89.2 vs 106 pmol/L [3.2 vs 3.8 ng/dL], *p* = 0.61).

### Prediction of Lateralizing PA using the 2025 Endocrine Society Criteria

According to the 2025 ES probability classifications, most patients 241/319 (75%), had an “intermediate” probability of having lateralizing PA, whereas 72/319 patients (23%) were categorized as “high” probability, and 6/319 (2%) as low probability (**Figure 2A**). As expected, baseline characteristics differed across these groups based on the defining features of the category. The high-probability group had lower potassium and PRA and higher PAC and ARR values (*p* <0.001 for all); however, blood pressure and number of antihypertensive medications were not significantly different (**Table 4**). Regardless of the probability classification, 61-78% of all patients were ultimately determined to have lateralizing PA in each group, with the high-probability group demonstrating the highest proportion of lateralizing disease (**Figure 2B**).

**Figure 2.**
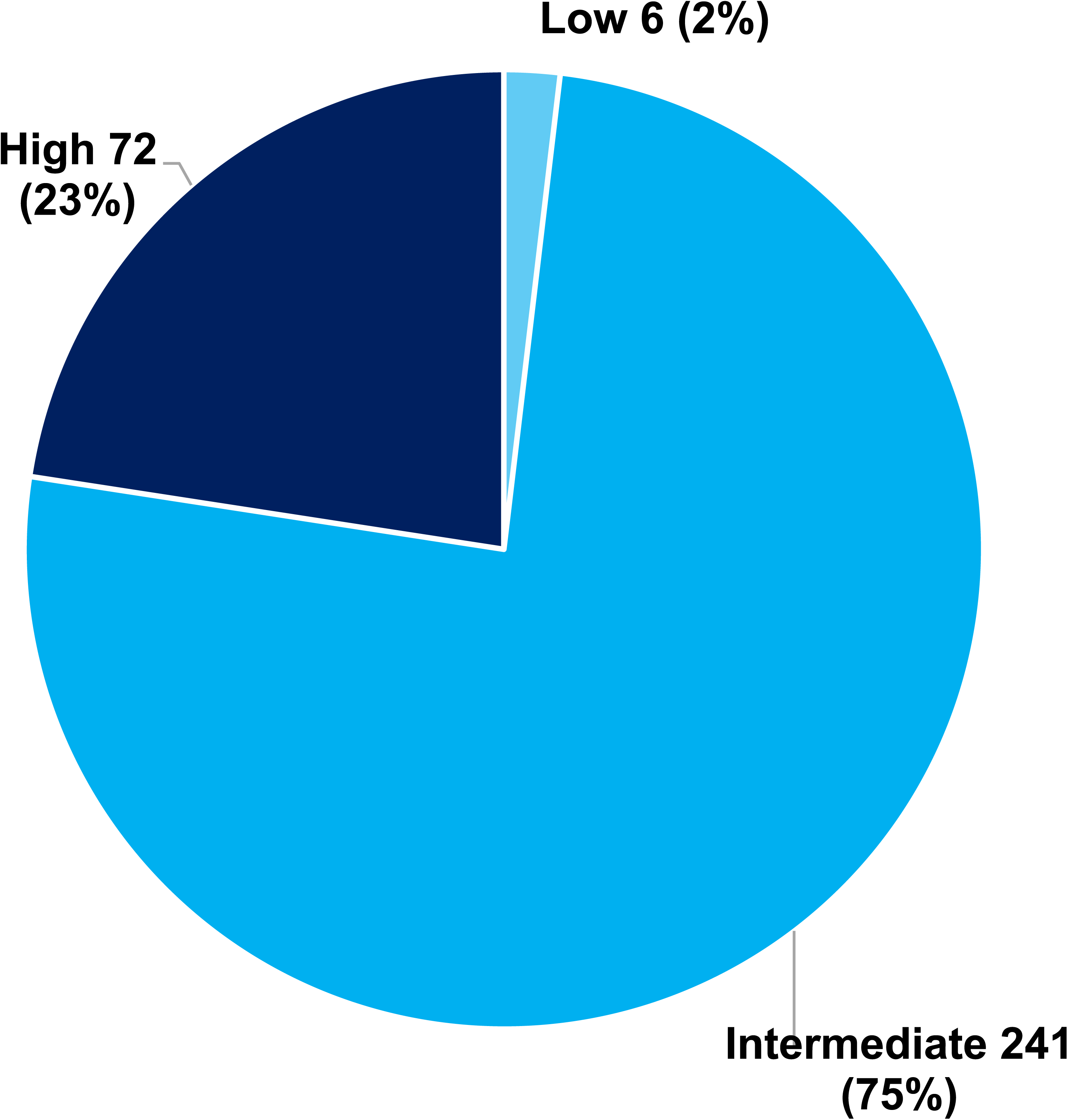
Distribution and Subtype Classification According to Probability of Lateralizing PA Based on the Endocrine Society 2025 Criteria (N = 319). (A) Distribution of patients according to the probability of lateralizing PA. (B) Subtype Classification According to Probability of Having Lateralizing PA. Abbreviation: PA, primary aldosteronism.

### Performance of the saline suppression test (SST) for predicting lateralizing PA

Mean serum potassium on the morning of SST was 3.86 ± 0.30 mmol/L. Hypokalemia (<3.5 mmol/L) at the time of SST was present in only three patients (1.6%), and potassium data were unavailable for one patient (0.5%). All four of these patients exhibited post-SST PAC levels above the diagnostic threshold (range: 250-646 pmol/L [9.0-23.3 ng/dL]); thus, there were no instances of hypokalemia resulting in a potentially false-negative result.

Overall, the vast majority of patients had a positive SST (86%, 162/188) whereas only a minority had a negative SST result (14%, 26/188). The proportion of PA patients with a positive SST was 50%, 85%, and 100% in the low-, intermediate-, and high-probability groups, respectively (**Figure 3**).

**Figure 3.**
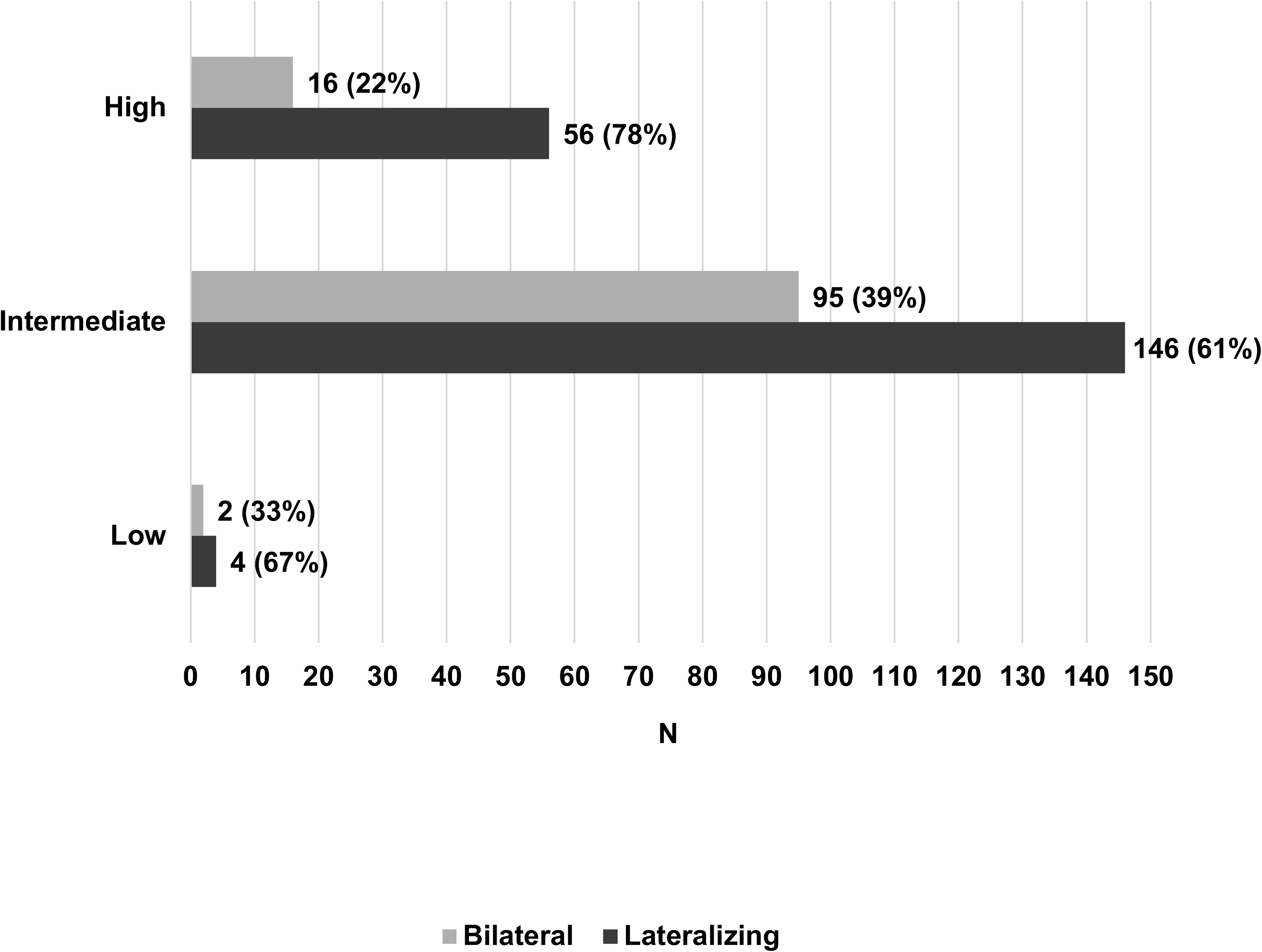
Probability of Lateralizing PA and SST Results (N= 188). The seated SST was considered positive if post-infusion PAC by immunoassay was≥ **217 pmol/L (**7.8 ng/dL), and negative if PAC fell below this threshold. Abbreviations: PA, Primary aldosteronism; SST, Saline suppress test; PAC, plasma aldosterone concentration

The discriminatory capacity of the SST to identify lateralizing PA was poor, as the majority of patients with both lateralizing and bilateral PA had positive SST results (**Figure 4A**; **Figure 5A**). Among 121 patients with confirmed lateralizing PA, 108 (89.3%) had a positive SST (i.e., true positive) and 13 (10.7%) had a negative SST (i.e., false negative). Conversely, among 67 patients with confirmed bilateral PA, 54 (80.6%) had a positive SST (i.e., false positive), 13 (19.4%) had a negative SST (i.e., true negative) (**Figure 4A**; **Figure 5A**). Accordingly, the SST demonstrated a sensitivity of 89.3% for detecting lateralizing PA but a specificity of 19.4% for excluding lateralizing PA. When evaluated with more granularity, the distribution of post-SST aldosterone levels exhibited near-complete overlap between those with lateralizing and non-lateralizing PA (**Figure 4B**), illustrating the limited discriminatory capacity of SST results for subtype differentiation regardless of the specific diagnostic threshold applied to the SST.

**Figure 4.**
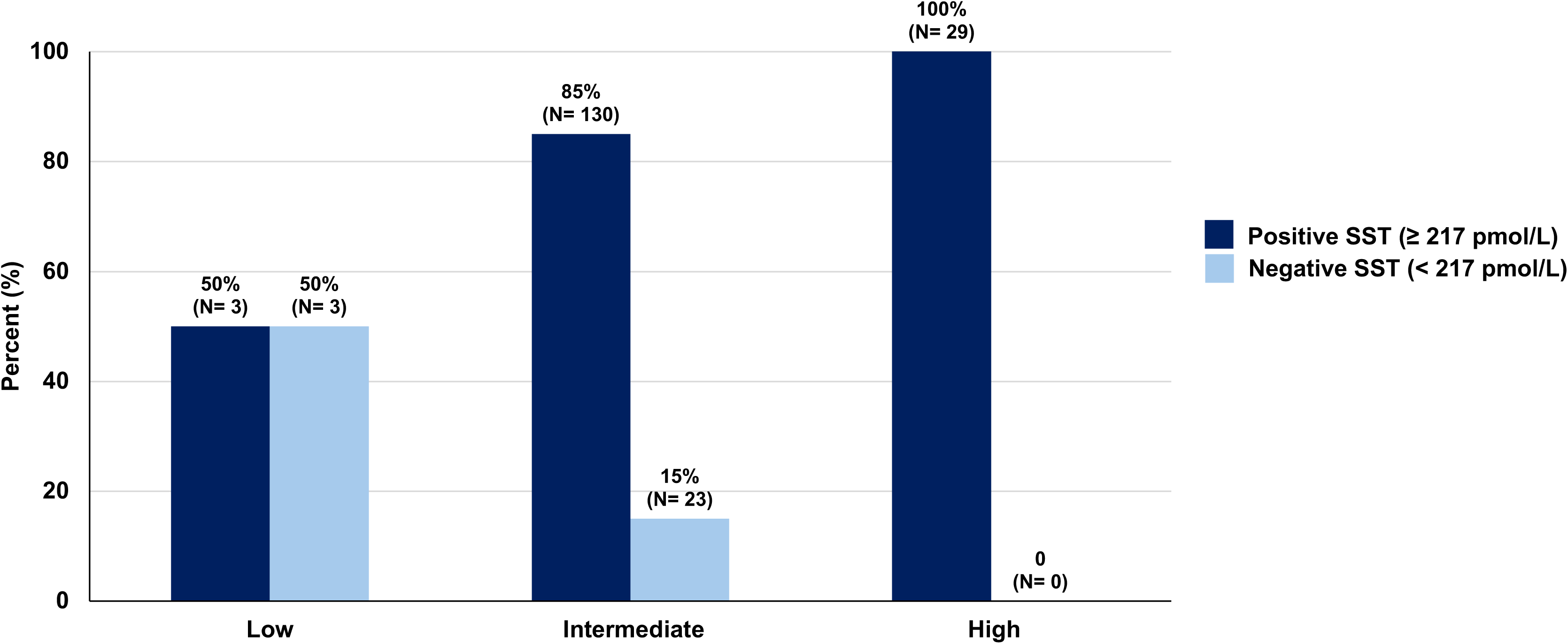
SST Results and Subtype Classification of PA (A) Distribution of SST Outcomes for all patients (N=188). (B) Post-SST Aldosterone Concentrations According to PA Subtype (N= 188). The seated SST was considered positive if post-infusion PAC by immunoassay was ≥ **217 pmol/L** (7.8 ng/dL), and negative if PAC fell below this threshold. The horizontal dashed line represents a post-SST aldosterone level of **217 pmol/L (**7.8 ng/dL). Each dot represents an individual patient, and horizontal lines indicate median values. Pairwise comparisons between subtypes were all statistically significant (Mann-Whitney U test, all p < 0.001). However, substantial overlap in post-SST PAC levels is observed between subtypes, demonstrating limited discriminatory ability of SST for subtype classification. Abbreviations: PA, Primary aldosteronism; SST, Saline suppression test.

**Figure 5.**
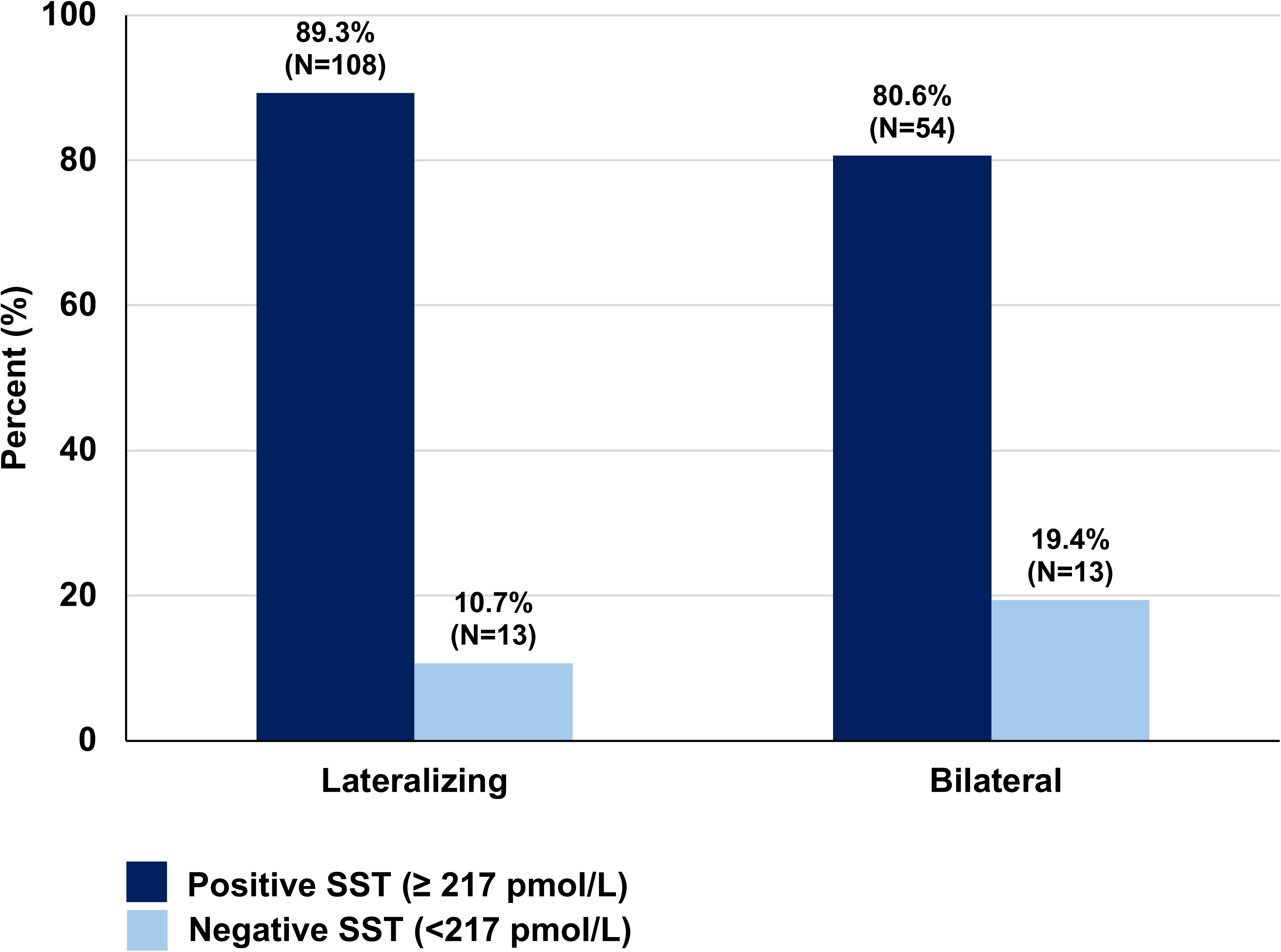
Distribution of SST Outcomes Across PA Subtypes. (**A) 2×2 Contingency Table for The Total Cohort (N= 188). (B) 2×2 Contingency Table for the Intermediate-Probability Group (n=153).** The seated SST was considered positive if post-infusion PAC by immunoassay was≥ **217 pmol/L** (7.8 ng/dL), and negative if PAC fell below this threshold. These tables illustrate the correlation between the seated SST results and PA subtype classification. Abbreviations: PA, Primary aldosteronism; SST, Saline suppression test.

This discordance and substantial overlap in post-SST aldosterone levels persisted even when focusing only on the “intermediate”-probability subgroup (**Figure 6A and 6B**), where the 2025 ES guidelines, and some expert opinions, specifically endorse performing this procedure to predict the likelihood of lateralizing PA and inform the use of imaging and AVS^4,24^. Again, the SST was positive in nearly all patients within the intermediate-probability subgroup, regardless of whether they ultimately had lateralizing or bilateral PA. Accordingly, while a positive SST was observed in 84 of 94 patients with lateralizing PA (true-positive = 89.4% and false-negative = 10.6%), 46 of 59 patients with bilateral PA also had a positive SST (false-positive = 78.0% and true-negative = 22.0%) (**Figure 5B and Figure 6A**). Thus, the sensitivity of the SST in predicting lateralizing PA in the intermediate probability group was 89.4% but with a specificity of 22.0%. By applying the ES approach and requiring SST as gatekeeper to AVS in intermediate probability, 23/153 (15%) would avoid AVS, of whom nearly half (10/23) would have had surgically amenable, lateralizing disease.

**Figure 6.**
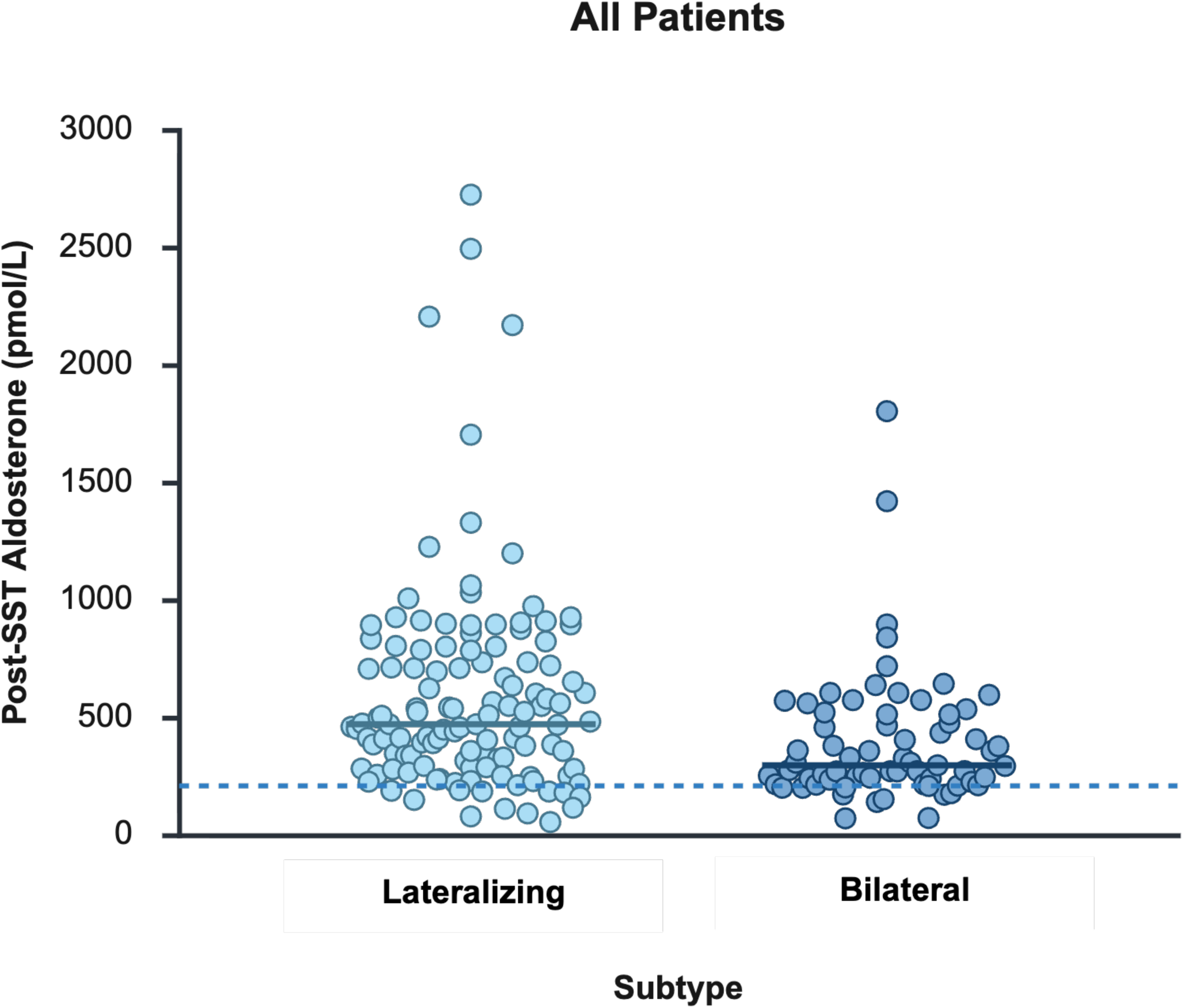
SST Results and Subtype Classification of PA Within the Intermediate-Probability Group (A) Distribution of SST Outcomes within the Intermediate-Probability Group (N=153). (B) Post-SST Aldosterone Concentrations According to PA Subtype in Patients with intermediate probability of having lateralizing PA (N= 153). The seated SST was considered positive if post-infusion PAC by immunoassay was ≥ 217 pmol/L (7.8 ng/dL), and negative if PAC fell below this threshold. The horizontal dashed line represents a post-SST aldosterone level of 217 pmol/L (7.8 ng/dL). Each dot represents an individual patient, and horizontal lines indicate median values. Pairwise comparisons between subtypes were all statistically significant (Mann-Whitney U test, all p < 0.001). However, substantial overlap in post-SST PAC levels is observed between subtypes, demonstrating limited discriminatory ability of SST for subtype classification. Abbreviations: PA, Primary aldosteronism; SST, Saline suppression test.

### Sensitivity Analyses

A sensitivity analysis was performed on the 47 patients with baseline serum potassium levels ≥ 4.0 mmol/L at the beginning of SST, rather than normokalemia (serum potassium ≥ 3.5 mmol/L), to more stringently exclude any potential PAC reductions by lower serum potassium concentrations. Even in this subgroup, most bilateral PA cases remained SST-positive, indicating persistent discordance between SST results and final subtype classification (**Table 5 and Figure 7**). Among 18 patients with bilateral PA, 17 had a positive SST (false-positives) and only 1 had a negative SST (true-negative). Among 29 patients with lateralizing PA, 24 had a positive SST (true-positives) and 5 had a negative SST (false-negatives).

**Figure 7.**
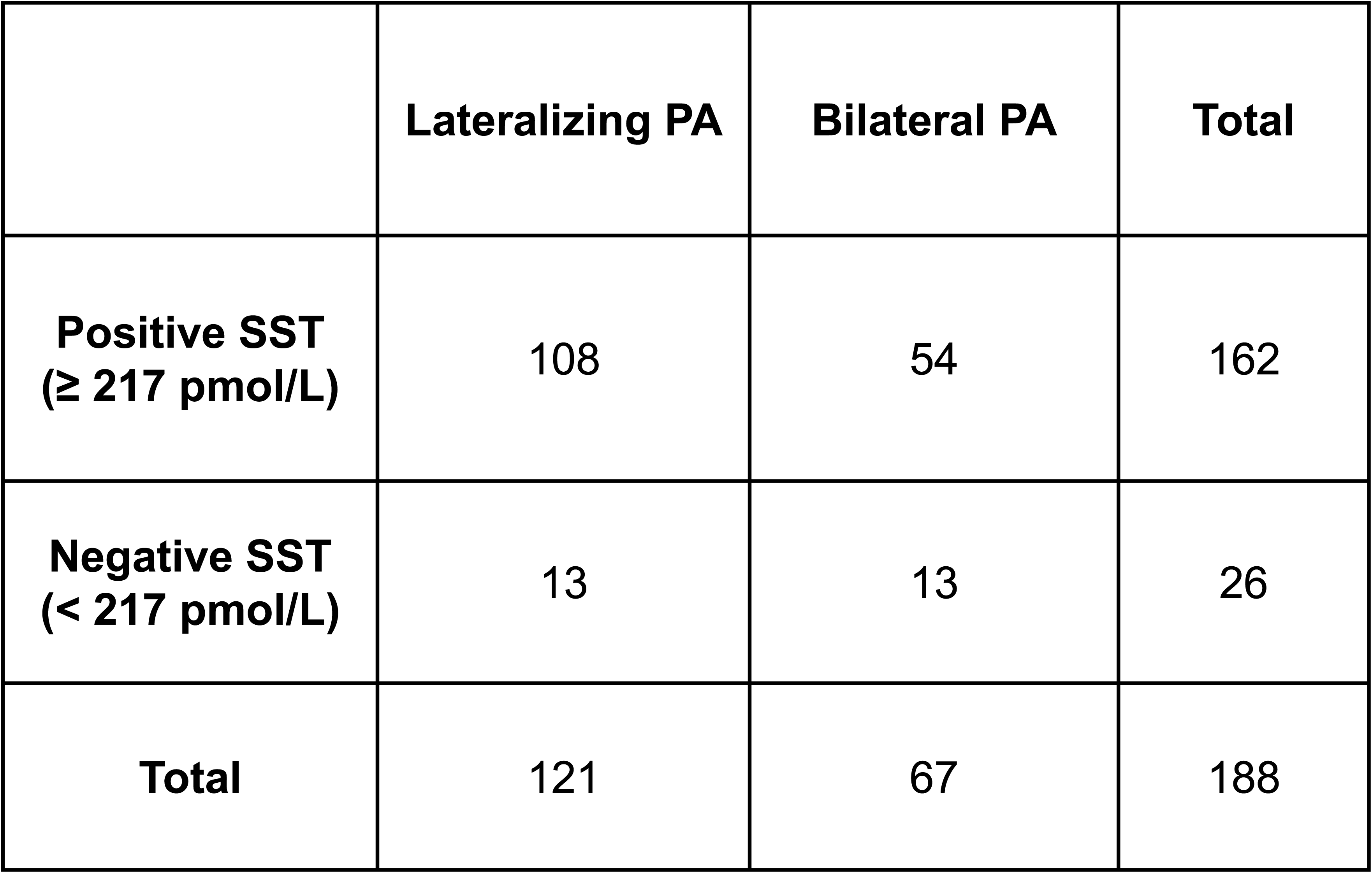
SST Results and Subtype Classification of PA in Patients with Baseline Serum Potassium Levels ≥ 4.0 mmol/L at the Beginning of SST and a 2×2 Contingency Table (n= 47). The seated SST was considered positive if post-infusion PAC by immunoassay was ≥ **217 pmol/L** (7.8 ng/dL), and negative if PAC fell below this threshold. Each dot represents an individual patient, and horizontal lines indicate median values. The horizontal dashed line represents a post-SST aldosterone level of **217 pmol/L (**7.8 ng/dL). Substantial overlap in post-SST PAC levels is observed between subtypes, demonstrating limited discriminatory ability of SST for subtype classification. Abbreviations: PA, Primary aldosteronism; SST, Saline suppression test.

**Table 5.**
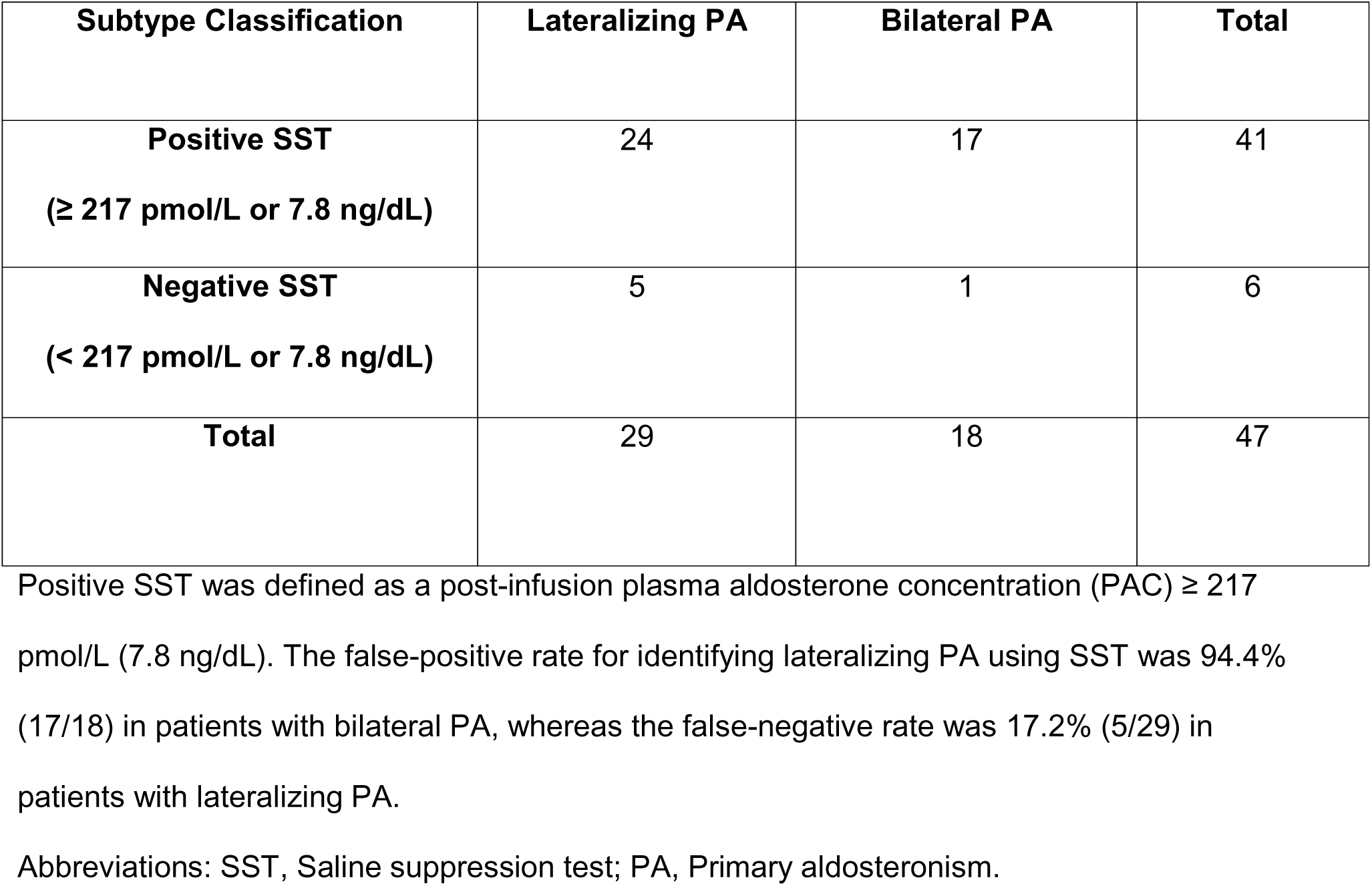
Diagnostic Performance of Seated SST in Subtyping PA in Patients with Baseline Serum Potassium Levels ≥ 4 mmol/L at the Beginning of SST (n=47)

Another sensitivity analysis was conducted to evaluate the diagnostic performance of the SST in differentiating PA subtypes for each method of determining subtype: AVS and PASO criteria following unilateral adrenalectomy. Again, the results of this sensitivity analysis echoed the same message as the main analyses. Among patients whose subtype was determined following unilateral adrenalectomy (n=87), the false-positive rate of SST was 90.9% whereas the false-negative rate was 10.5% (**Table 6**). Similarly, among patients whose subtype was determined using AVS (n=122), the false-positive rate of SST was 80.0% and the false-negative rate was 9.7% (**Table 7**).

**Table 6.** Diagnostic Performance of Seated SST in Differentiating PA Subtypes in Surgery-Confirmed Cohorts (n= 87)

| **Surgery-Confirmed** | **Lateralizing PA** | **Bilateral PA** | **Total** |
| --- | --- | --- | --- |
| **Positive SST** | 68 | 10 | 78 |
| **Negative SST** | 8 | 1 | 9 |
| **Total** | 76 | 11 | 87 |

Positive SST was defined as a post-infusion PAC ≥ 217 pmol/L (7.8 ng/dL). The false-positive rate for identifying lateralizing PA using SST was 90.9% (10/11) in patients with bilateral PA, whereas the false-negative rate was 10.5% (8/76) in patients with lateralizing PA.
Abbreviations: SST, Saline suppression test; AVS, Adrenal venous sampling; PA, Primary aldosteronism.

**Table 7.** Diagnostic Performance of Seated SST in Differentiating PA Subtypes in AVS-Confirmed Cohorts (n= 122)

| **AVS-Confirmed** | **Lateralizing PA** | **Bilateral PA** | **Total** |
| --- | --- | --- | --- |
| **Positive SST** | 56 | 48 | 104 |
| **Negative SST** | 6 | 12 | 18 |
| **Total** | 62 | 60 | 122 |

Positive SST was defined as a post-infusion PAC ≥ 217 pmol/L (7.8 ng/dL). The false-positive rate for identifying lateralizing PA using SST was 80.0% (48/60) in patients with bilateral PA, whereas the false-negative rate was 9.7% (6/62) in patients with lateralizing PA.
Abbreviations: SST, Saline suppression test; AVS, Adrenal venous sampling; PA, Primary aldosteronism

Since 53 patients had unilateral adrenalectomy prior to the publication of the PASO criteria, and thus did not have complete information to assess post-operative biochemical outcomes, sensitivity analyses were conducted after excluding these 53 individuals (remaining sample size n=266). Of note, these excluded individuals almost certainly had either lateralizing PA and/or remarkable clinical improvements following unilateral adrenalectomy (**Table 8**), thus justifying the operative decision. Excluding them in this sensitivity analysis was performed only to demonstrate consistency and compliance with PASO-style reporting, which has become the current research standard. Following this exclusion, the probability classifications once again showed that most patients fell in the “intermediate” category, and that regardless of the probability classification, 50-76% of all patients were ultimately determined to have lateralizing PA (**Figure 8A and 8B**). The number of patients with available SST information declined to 159 (from 188); however, the discriminatory capacity of the SST to predict subtyping outcomes remained unchanged and poor: the SST was “positive” the vast majority of the time (**Figure 9**), resulting in a sensitivity and specificity were 89.2% and 19.7%, respectively, with a 80.3% false-positive rate and 10.8% false-negative rate (**Figure 10A**). The discriminatory capacity of the SST was again similarly poor when restricted to only those with an “intermediate” probability for lateralizing PA: sensitivity = 86.6%, specificity = 21.7%, false-positive rate = 78.3%, false-negative rate = 13.4% (**Figure 10B**).

**Figure 8.**
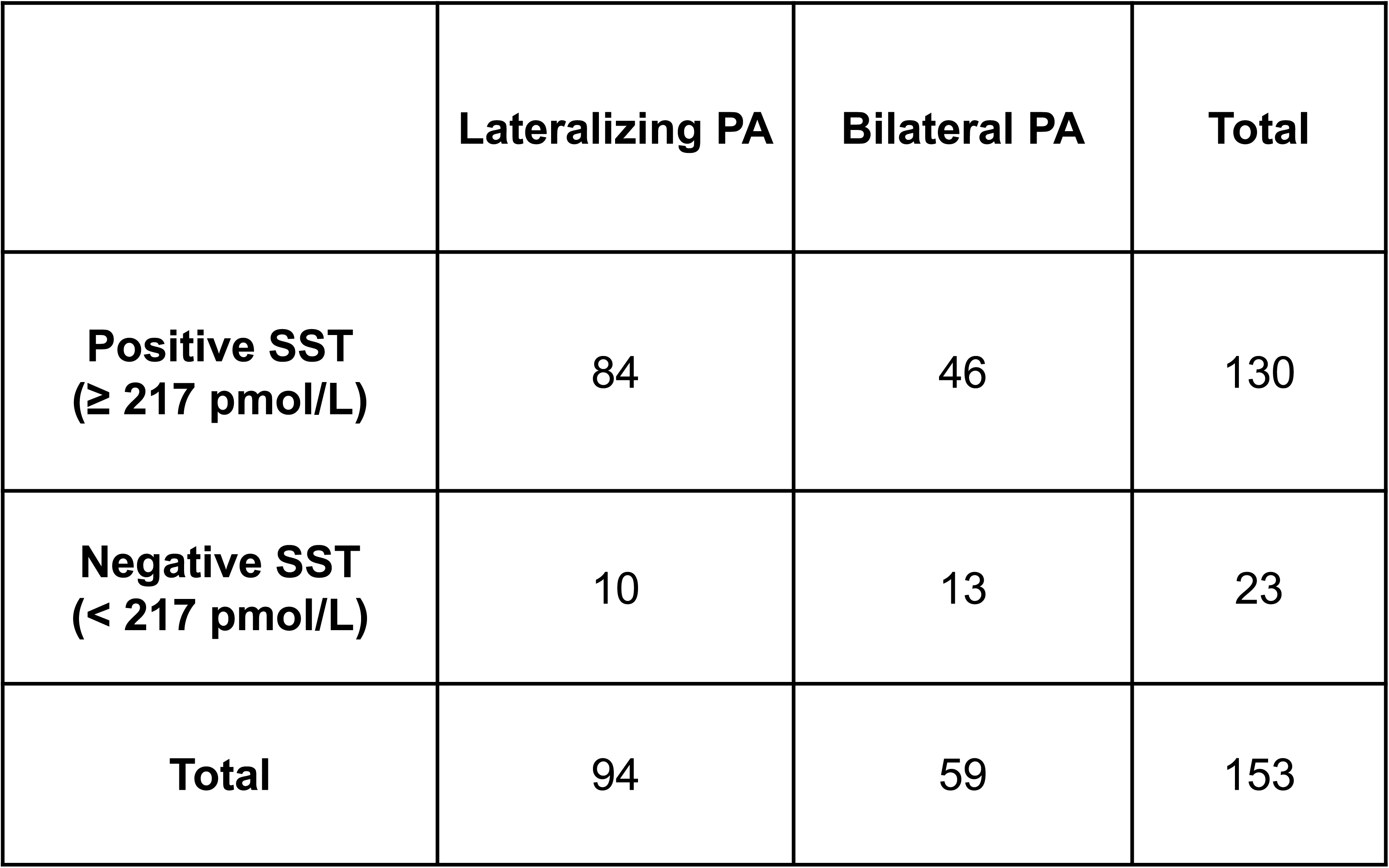
Distribution and Subtype Classification According to Probability of Lateralizing PA Based on the Endocrine Society 2025 Criteria, Excluding Those Without Post-Operative Outcomes (N = 266). (A) Distribution of patients according to the probability of lateralizing PA. (B) Subtype Classification According to Probability of Having Lateralizing PA. Abbreviation: PA, primary aldosteronism

**Figure 9.**
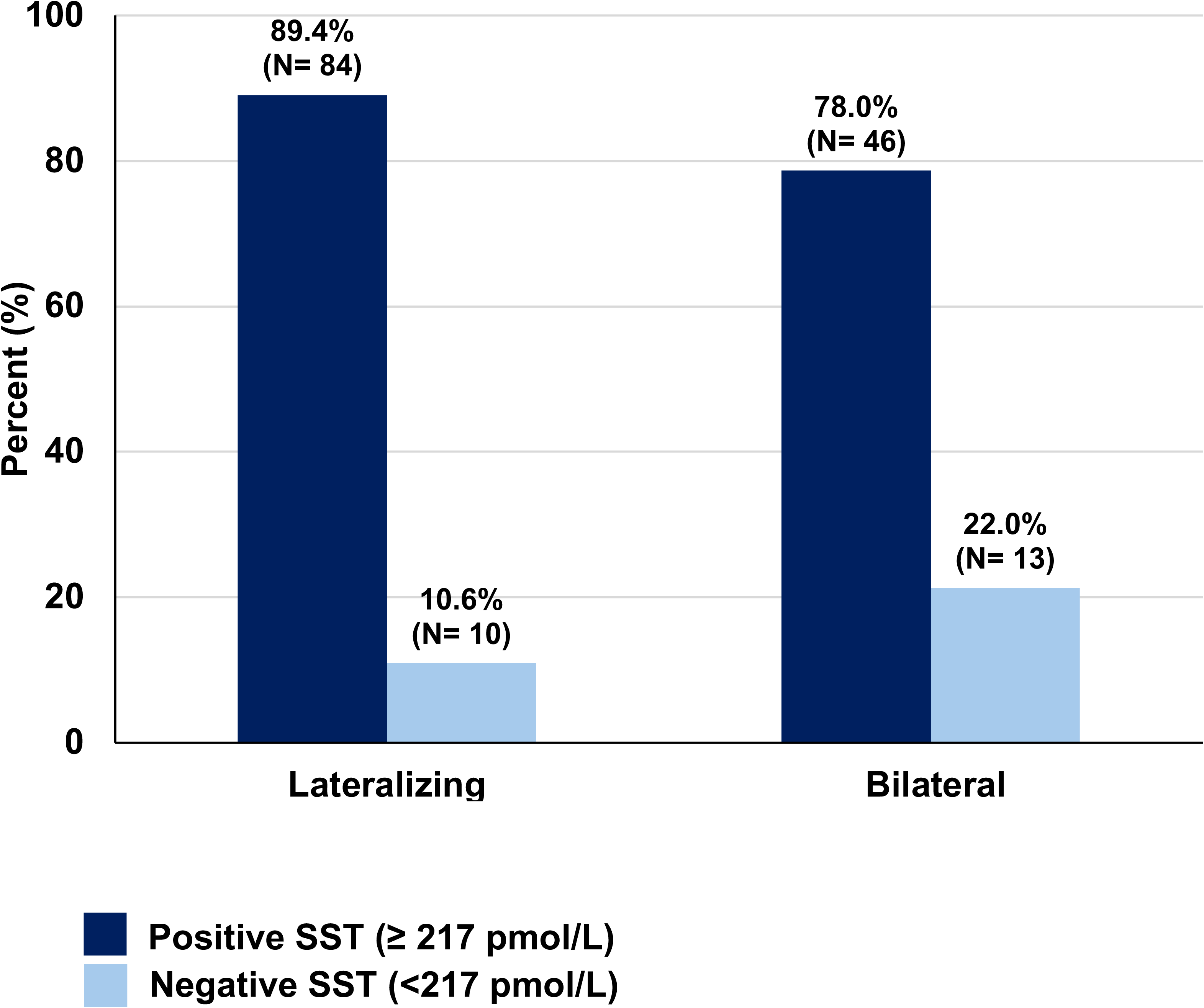
Probability of Lateralizing PA and SST Results, Excluding Those Without Postoperative Outcomes (N= 159). The seated SST was considered positive if post-infusion PAC by immunoassay was≥ **217 pmol/L** (7.8 ng/dL), and negative if PAC fell below this threshold. Abbreviations: PA, Primary aldosteronism; SST, Saline suppress test; PAC, plasma aldosterone concentration

**Figure 10.**
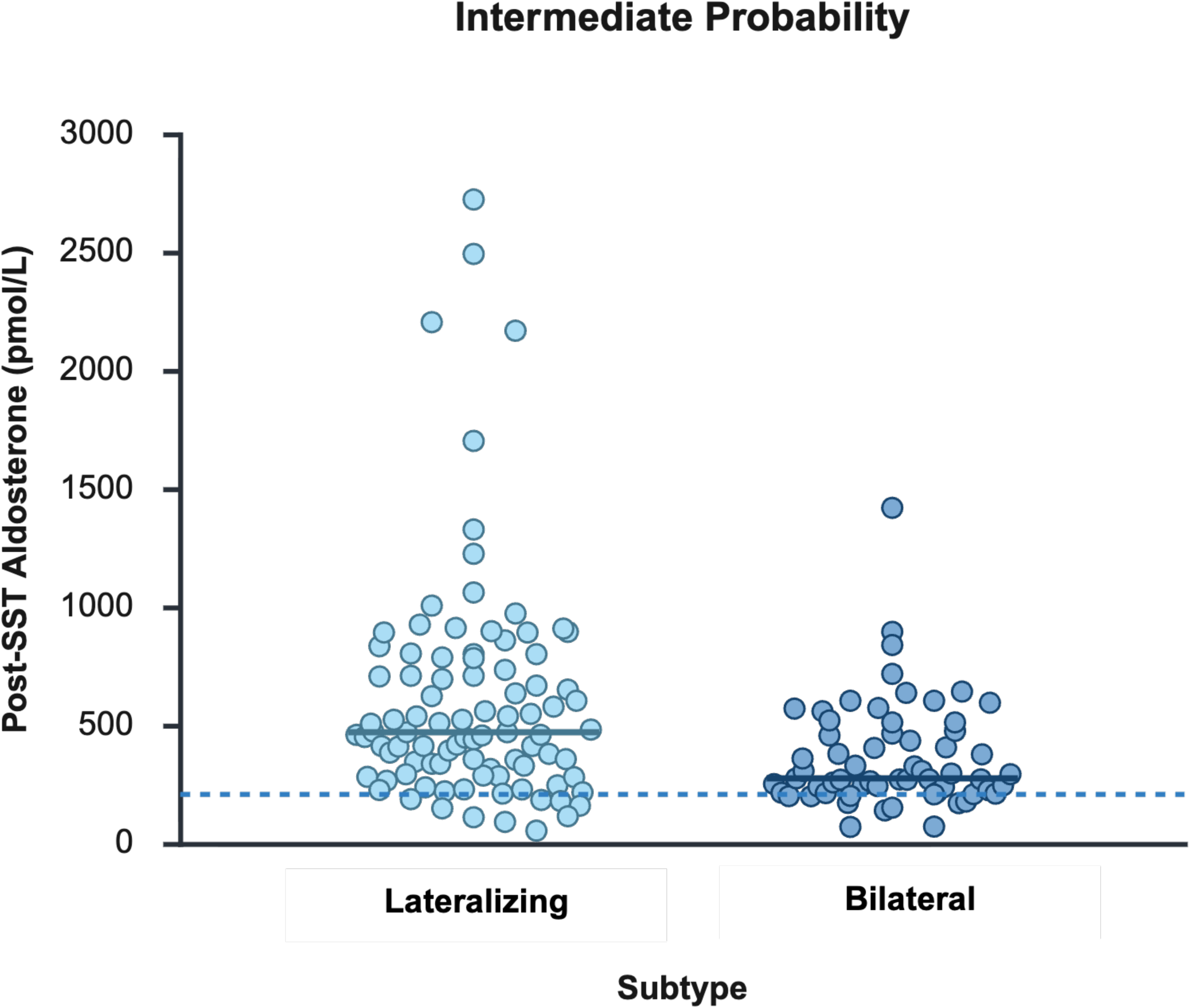
SST Results and Subtype Classification of PA, Excluding Patients Without Post-Operative Outcomes. (A) Post-SST Aldosterone Concentrations According to PA Subtype and 2×2 Contingency Table (N= 159). (B) Post-SST Aldosterone Concentrations According to PA Subtype in Patients with Intermediate Probability of having Lateralizing PA and 2×2 Contingency Table (N= 127). The seated SST was considered positive if post-infusion PAC by immunoassay was ≥ **217 pmol/L** (7.8 ng/dL), and negative if PAC fell below this threshold. Each dot represents an individual patient, and horizontal lines indicate median values. The horizontal dashed line represents a post-SST aldosterone level of **217 pmol/L** (7.8 ng/dL). Substantial overlap in post-SST PAC levels is observed between subtypes, demonstrating limited discriminatory ability of SST for subtype classification. Abbreviations: PA, Primary aldosteronism; SST, Saline suppression test.

**Table 8.**
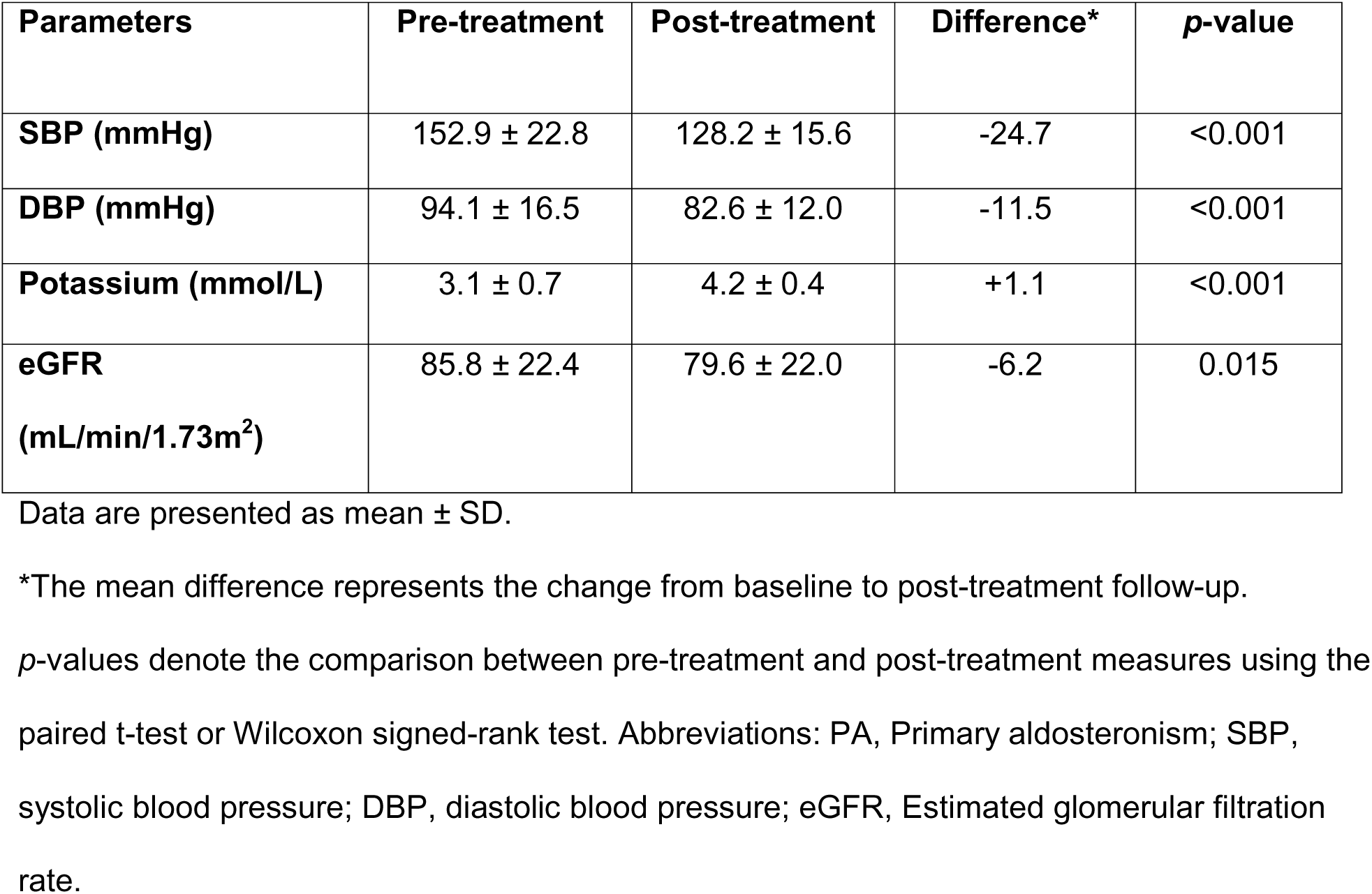
Treatment Outcomes of Unilateral Adrenalectomy for PA (n= 50)

| Parameters | Pre-treatment | Post-treatment | Difference* | *p*-value |
| --- | --- | --- | --- | --- |
| **SBP (mmHg)** | 152.9 ± 22.8 | 128.2 ± 15.6 | -24.7 | <0.001 |
| **DBP (mmHg)** | 94.1 ± 16.5 | 82.6 ± 12.0 | -11.5 | <0.001 |
| **Potassium (mmol/L)** | 3.1 ± 0.7 | 4.2 ± 0.4 | +1.1 | <0.001 |
| **eGFR**<br>**(mL/min/1.73m<sup>2</sup>)** | 85.8 ± 22.4 | 79.6 ± 22.0 | -6.2 | 0.015 |

Data are presented as mean ± SD.
\*The mean difference represents the change from baseline to post-treatment follow-up.
*p*-values denote the comparison between pre-treatment and post-treatment measures using the paired t-test or Wilcoxon signed-rank test. Abbreviations: PA, Primary aldosteronism; SBP, systolic blood pressure; DBP, diastolic blood pressure; eGFR, Estimated glomerular filtration rate.

Additional sensitivity analyses were performed to evaluate the diagnostic performance across different AVS techniques and demonstrated that the AVS method did not impact the results (**Table 9**). The sensitivity of AVS for detecting lateralizing PA based on surgical outcomes remained consistently high at 91.4% when AVS was performed with ACTH stimulation (n=144) and 100% when AVS was performed without ACTH stimulation (n=39). In addition, when using a stricter LI ≥4 threshold (n= 106), the results remained unchanged.

**Table 9.** Sensitivity Analysis of Differing AVS Techniques.

| **AVS** | **n** | **Sensitivity (%)** | **Specificity (%)** |
| --- | --- | --- | --- |
| **ACTH-stimulated**<br>**AVS** | 144 | 91.4 | 95.9 |
| **Non-stimulated AVS** | 39 | 100 | 88.9 |

The ACTH-stimulated AVS group includes patients from King Chulalongkorn Memorial Hospital (2020–2021) and Phramongkutklao Hospital (2016–2025). The non-stimulated AVS group includes patients from King Chulalongkorn Memorial Hospital who underwent the procedure after 2021 (2021-2025). The sensitivity and specificity were calculated based on patients with successful cannulation and a confirmed final diagnosis of either lateralizing or bilateral PA based on postoperative outcomes.
Abbreviations: AVS, adrenal venous sampling; n, number of patients; PA, primary aldosteronism.

When additional sensitivity analyses were done to evaluate alternative interpretations for "intermediate" probability for lateralizing PA, defined as patients with PAC levels between 305-554 pmol/L (11–20 ng/dL) and normokalemia (serum potassium ≥ 3.5 mmol/L) without a history of hypokalemia, or those having PAC levels within this range regardless of serum potassium, as well as patients presenting without hypokalemia, the findings remained essentially unchanged (**Table 10-12**).

**Table 10.** Diagnostic Performance of Seated SST in Subtyping PA in Patients Within the Intermediate Probability Group (PAC levels between 305-554 pmol/L [11–20 ng/dL]) Who Had Concurrent Normokalemia (Serum Potassium ≥ 3.5 mmol/L) and No History of Hypokalemia (N= 18). The seated SST was considered positive if post-infusion PAC by immunoassay was ≥ 217 pmol/L (7.8 ng/dL), and negative if PAC fell below this threshold. Abbreviations: PA, Primary aldosteronism; SST, Saline suppression test; PAC, plasma aldosterone concentration

|  | Lateralizing PA | Bilateral PA | Total |
| --- | --- | --- | --- |
| **Positive SST (≥ 217 pmol/L)** | 10 | 3 | 13 |
| **Negative SST (< 217 pmol/L)** | 2 | 3 | 5 |
| **Total** | 12 | 6 | 18 |

Sensitivity, 83.3%; Specificity, 50.0 %; False-positive rate, 50.0%; False-negative rate 16.7%

**Table 11.**
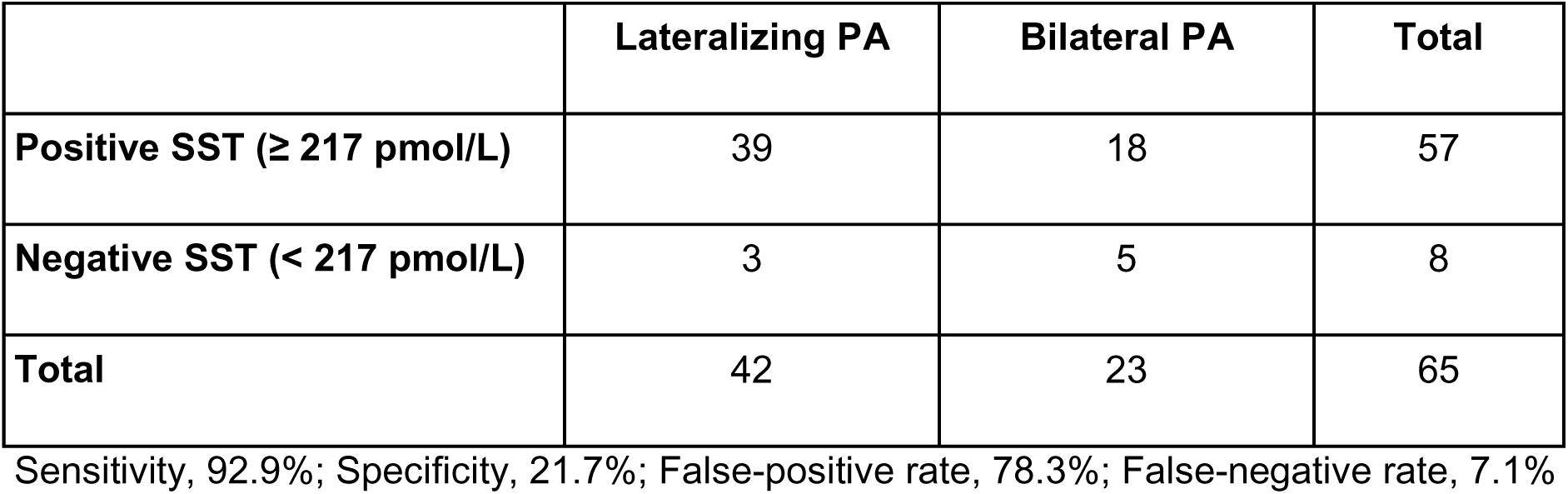
Diagnostic Performance of Seated SST in Subtyping PA in Patients Within the Intermediate Probability Group (PAC levels between 305-554 pmol/L [11–20 ng/dL]) Regardless of Potassium Status (n= 65). The seated SST was considered positive if post-infusion PAC by immunoassay was ≥ 217 pmol/L (7.8 ng/dL), and negative if PAC fell below this threshold. Abbreviations: PA, Primary aldosteronism; SST, Saline suppression test; PAC, plasma aldosterone concentration

|  | Lateralizing PA | Bilateral PA | Total |
| --- | --- | --- | --- |
| **Positive SST ($\geq 217$ pmol/L)** | 39 | 18 | 57 |
| **Negative SST ($< 217$ pmol/L)** | 3 | 5 | 8 |
| **Total** | 42 | 23 | 65 |

Sensitivity, 92.9%; Specificity, 21.7%; False-positive rate, 78.3%; False-negative rate, 7.1%

**Table 12.**
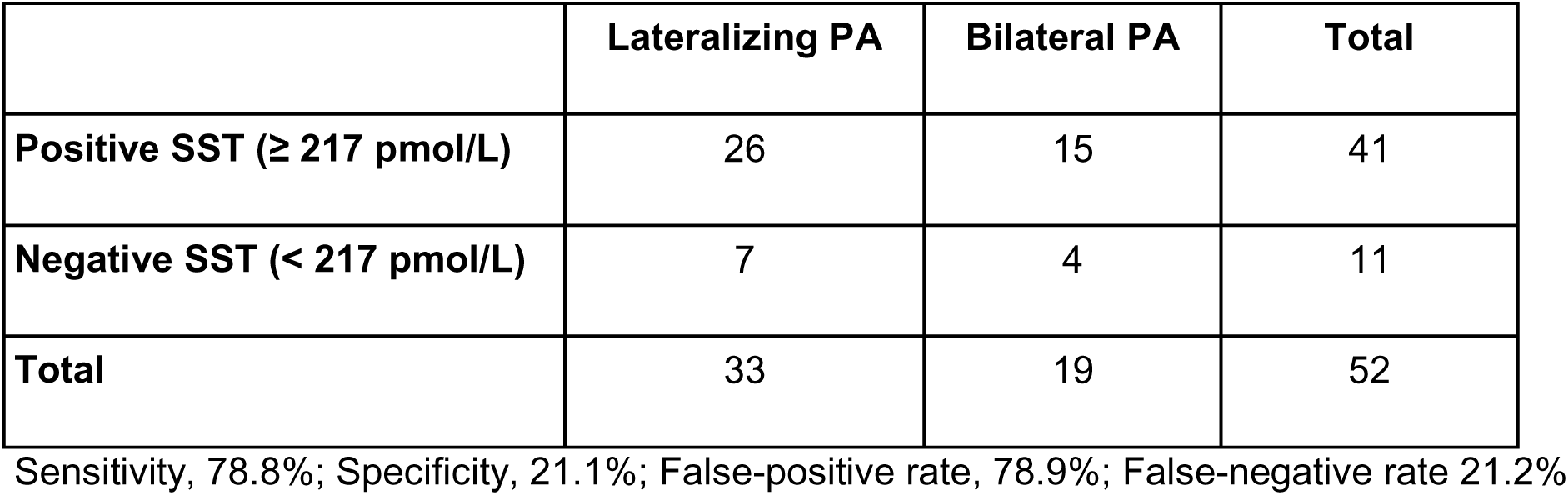
Diagnostic Performance of Seated SST in Subtyping PA in Patients Who Presented With Normokalemia (Serum Potassium ≥ 3.5 mmol/L) (n= 52). The seated SST was considered positive if post-infusion PAC by immunoassay was ≥ 217 pmol/L (7.8 ng/dL), and negative if PAC fell below this threshold. Abbreviations: PA, Primary aldosteronism; SST, Saline suppression test.

|  | Lateralizing PA | Bilateral PA | Total |
| --- | --- | --- | --- |
| **Positive SST ($\geq 217$ pmol/L)** | 26 | 15 | 41 |
| **Negative SST ($< 217$ pmol/L)** | 7 | 4 | 11 |
| **Total** | 33 | 19 | 52 |

Sensitivity, 78.8%; Specificity, 21.1%; False-positive rate, 78.9%; False-negative rate 21.2%

## Discussion

This study evaluated the performance of the 2025 ES probability framework for subtype differentiation, and the utility of aldosterone suppression testing for predicting PA subtype, in a large cohort of patients with PA at two specialized referral centers. Though observational studies are susceptible to bias and confounding, two key strengths of this study are the fact that the majority of patients were ultimately found to have lateralizing PA even though most were classified as having an “intermediate” probability of having lateralizing PA, and that Thai clinicians at these study sites followed traditional international guideline practices for testing and aldosterone suppression testing, yet also routinely overruled the results of these outcomes and pursued subtyping ascertainment regardless. Because of these unique features, the analyses in this study were able to assess the performance of probability classifications in predicting outcomes, and the accuracy of aldosterone suppression test results in predicting subtypes, even when the SST was “negative”. The findings generally supported the recent ES guideline recommendation that aldosterone suppression testing can be skipped in patients with a “high” probability of having lateralizing PA. However, beyond this subgroup, reliance on guideline-endorsed probability classifications alone was not useful for predicting PA subtype, as the frequency of lateralizing PA was similar across probability designations. Even though the sample size of patients in the “low” probability group was small, the misclassification rate of lateralizing PA was marked. In the intermediate probability group, where guidelines endorse using aldosterone suppression testing to determine the need for AVS, using the SST to identify those who would benefit from AVS^4,24^ also performed poorly, with unacceptable false-positive rates (>80% of people with bilateral PA had a positive SST) and false-negative rates (>10% of people with lateralizing PA had a negative SST). These results were reproduced following multiple sensitivity analyses to interrogate their durability. This lack of discriminatory capacity was primarily driven by the fact that the majority of patients had a positive SST result, regardless of whether their ultimate subtype was lateralizing or bilateral PA, highlighting the limited utility of aldosterone suppression testing as an individualized triage tool.

Our results build on the growing number of studies questioning the role of aldosterone suppression testing in the evaluation of PA^15–18,25^. A systematic review and meta-analysis of all major studies that evaluated aldosterone suppression tests concluded that the use of these tests to “confirm” PA was based on very low-quality evidence that was marred by high risk for bias, over-estimation of value, and the general lack of reference standards by which to ensure confidence in the veracity of the results^25^. This general conclusion was further amplified by the systematic review and meta-analysis conducted by the Endocrine Society to accompany the 2025 guidelines, where only very low-quality evidence for the use of aldosterone suppression testing could be identified, and was therefore insufficient to continue recommending its use for diagnostic purposes^13^. Finally, in the first prospective and blinded study to evaluate the diagnostic performance of the SST in predicting treatment outcomes, Leung et al. showed that the SST result could not meaningfully predict treatment responses to targeted therapy, and frequently misclassified patients such that the diagnosis of PA and targeted therapy could have been erroneously missed altogether^16^. These comprehensive and rigorous assessments prompted the 2025 ES guideline update to no longer recommend aldosterone suppression testing for the purposes of diagnosing (or excluding) PA. While this was a major update that could have simplified the diagnostic approach by removing unvalidated practices, it was also accompanied by a new suggestion that aldosterone suppression testing may instead be used in individuals with an “intermediate” probability of lateralizing PA to help triage the use of AVS resources. As seen in our study, the vast majority of PA patients fell into the “intermediate” probability classification, thus the main repercussion of the new ES guidance is a shift in the use of aldosterone suppression testing from an inaccurate diagnostic to an unvalidated predictor of lateralization in almost all patients with PA. The problem with this evidence-free endorsement has been elegantly articulated by Demko and Cohen^26^, *“…confirmatory testing was originally intended to establish the presence of autonomous aldosterone production, not to determine laterality…when a test designed to establish disease presence is repurposed to predict disease subtype, poor discrimination may be a signal that the test is being asked to answer a question it was never intended to address.”*

It should be noted that some believe that a high sensitivity is a virtue of an aldosterone suppression test for predicting PA subtype. However, when the purpose of the test is gatekeeping to prevent unnecessary procedures, high sensitivity is neither the goal nor the measure of success. As we observed in this study and as has been shown in prior studies^17,18^, the inability of SST to meaningfully discriminate between bilateral and lateralizing cases undermines any value of the test as a gatekeeper for invasive subtyping procedures. Leung et al. showed that of the small minority of patients who would have been spared an AVS, half had lateralizing disease that would have been missed, and conversely, nearly as many patients with bilateral disease would have undergone unnecessary AVS as if no SST were performed to guide triage^17^. Similarly, Tsai et al. showed that even if the most liberalized criteria for the SST were used to maximize sensitivity and minimize false-negative predictions, this rubric resulted in nearly half of patients with bilateral disease having a false-positive test, resulting in unnecessary AVS^18^. This latter phenomenon is essentially replicated in our current study, where post-SST PAC levels overlap so extensively in those with and without lateralizing PA that any attempt to distinguish PA subtypes with a single, static threshold yields unacceptable accuracy and erroneous interpretations. The failure of the SST to make this distinction should not come as a surprise, as the regulation of aldosterone in PA depends on many more factors than acute changes in volume and renin-angiotensin II biology, many of which are unaccounted for when performing the SST (and other common aldosterone suppression tests^27–31^).

Crucially, our study demonstrated that SST has limited ability to discriminate between lateralizing and bilateral PA, both in the whole cohort and the intermediate-probability group as defined by the 2025 ES Guideline. In our cohort, the "positive" rate of the seated SST was nearly identical across the intermediate- and high-probability categories, indicating that enforcing this step in the treatment pathway may provide no additional diagnostic refinement or triage value. Under the proposed guideline algorithms, this non-refining diagnostic step would be mandated for approximately 75% of all PA patients, imposing an immense and unnecessary procedural burden. As a result, few AVS procedures are safely avoided, while a meaningful proportion of true lateralizing cases would be inadvertently missed among those blocked by having "negative" SST results. Beyond diagnostic performance, resource constraints in many healthcare systems further limit access to the aldosterone suppression test^21^, making it even more critical that any mandatory preceding test provides meaningful clinical value. Enforcing a gatekeeper test that lacks clinical granularity subjects patients to unnecessary procedures and hinders access to definitive treatment, particularly in resource-limited settings.

While various predictive models have been proposed to identify lateralizing disease without AVS, their performance also remains inconsistent across external validations^32–35^. Studies using post-SST aldosterone cutoffs have reported that a PAC threshold of 13.1 ng/dL achieved a sensitivity of 78–94% and specificity of 79–95% in predicting lateralizing disease in Japanese cohorts^36,37^. However, when using the same threshold, another study reported lower performance, with a sensitivity of 68% and specificity of 57%, highlighting limited reproducibility^38^. Recently, an expert consensus proposed a clinical severity score incorporating blood pressure, basal PAC, and serum potassium levels to stratify patients into “mild,” “moderate,” and “severe” PA^39^. This framework aimed to prioritize patients for AVS in a resource-conscious manner, and indeed, the proportion of patients with lateralizing disease increased with greater clinical severity. However, despite this graded association, the score did not provide sufficient accuracy for definitive subtype classification at the individual level, reinforcing the continued need for AVS in patients being considered for surgery. Other non-invasive methods to predict PA subtype, including machine learning models with or without integration of steroid profiling^34,40–42^, have been proposed to refine patient selection for targeted surgical therapy. Emerging nuclear imaging techniques also show promise; however, these approaches are currently hampered by limited availability, inconsistent validation across diverse populations^43–46^, and high costs.

It should be noted that measurements of PAC in our current study were performed using an immunoassay, whereas the gold-standard method for measuring any adrenocortical steroids is via liquid chromatography with tandem mass spectroscopy (LC-MS/MS). However, because the vast majority of worldwide clinicians use immunoassays to measure PAC, and LC–MS/MS is not yet widely accessible in many parts of the world, international guidelines primarily recommend immunoassay-based PAC measurements, with specific guidance for immunoassay interpretations, which we followed exactly in this study^4^. In addition, diagnostic accuracy studies have shown that no PAC threshold reliably predicts treatment responses or subtyping results^16,17,47^, regardless of the assay used. In addition, the 2025 ES probability framework does not account for intraindividual variability of aldosterone levels^48–50^. Even among patients with overt PA, it is not uncommon that PAC measurements may fall below diagnostic thresholds^48^. Consequently, on any given day of assessment, patients with truly “high”-risk disease may frequently be classified as “intermediate”, and vice versa. This variability of aldosterone production and phenotype (seen with both immunoassay and LC-MS/MS measurements)^48,50,51^ may partially explain why the proportions of lateralizing PA in our study were comparable between groups, regardless of their initial probability designation. A similar limitation applies to post-SST aldosterone measurements, as patients with lateralizing disease may still demonstrate apparent aldosterone suppression following the test^14–18^, further reducing the reliability of these approaches for subtype classification. One may argue that many patients categorized as having “intermediate” probability in our cohort actually had a “high” probability of PA (based on their aldosterone and potassium values), even though we applied the ES guideline definitions for these categories exactly as written. To address this, we performed multiple sensitivity analyses, including stricter definitions of the intermediate probability, and the results remained unchanged.

Our study has limitations. Observational and retrospective studies can suffer from residual confounding and bias. Undoubtedly, patient selection and test performance in this study were influenced by clinical decisions that were not always standardized; however, the strength of this study relied on the fact that Thai clinicians at these high-volume PA centers routinely performed subtyping procedures regardless of initial screening results and regardless of SST outcomes. In addition, while multicenter studies are often limited by inter-laboratory variability even when the same assays are used, all hormonal measurements in this study, including aldosterone and renin, were performed in a single laboratory, ensuring analytical consistency. By leveraging this approach, we were able to assess the discriminatory capacity of the ES probability framework and the SST for predicting PA subtype. While it is possible that the point estimates we observed may have been different had this study been conducted in a prospective protocol, the overlap in the post-SST results between lateralizing and bilateral PA, and the consistency of these results with prior prospective and retrospective studies from different countries^17,18^, provides reassurance for the validity and reproducibility of the results. A second limitation is that a slight majority of the PA patients in this study had lateralizing PA, reflecting diagnostic and referral biases that resulted in over-representation of lateralizing PA and an under-representation of bilateral PA. However, this feature proved valuable as it ensured an ample population of individuals with lateralizing PA with which to test the accuracy of the probability classifications and SST results designed to predict this very subtype. In addition, a selection bias must be acknowledged in the pre-operative workflow, as the decision to disregard the seated SST results and proceed directly to AVS was heavily guided by baseline clinical severity and a high pre-test probability of having lateralizing PA, which could have resulted in an overestimation of false-negative rates. However, sensitivity analyses restricted to those with intermediate PAC levels and no history of hypokalemia, representing a milder clinical phenotype (**Supplemental Table 8**), still yielded a false-negative rate of 16.7%, suggesting that this limitation did not fully account for the observed diagnostic gaps. A fourth limitation is that not all patients in our cohort underwent AVS, and surgical outcomes alone were used as a reference standard for some patients^20^. However, this approach aligns with clinical guidelines and practices^4,12,19,22^, which state that patients with severe PA with a unilateral adrenal lesion can skip AVS and proceed directly to unilateral adrenalectomy. To ensure that these patients did not skew our results, we performed sensitivity analyses in the AVS-confirmed and surgery-confirmed cohorts and showed similar outcomes, supporting the robustness of our reference standard. Additionally, potential diagnostic variations arising from AVS technical heterogeneity should be noted; as demonstrated in **Supplemental Table 7**, a slightly lower specificity was observed in non-stimulated AVS compared to ACTH-stimulated protocols, suggesting a potential susceptibility to false-positive lateralization in non-stimulated cohorts that warrants cautious clinical interpretation during surgical selection. Fifth, a minority of patients who underwent unilateral adrenalectomy prior to 2018 had PA subtype classification based on clinical outcomes alone. We acknowledge that patients with bilateral PA may also experience clinical improvement^23,52–54^, potentially leading to misclassification. However, sensitivity analyses excluding these patients reproduced the results of the primary analysis. In addition, our previously published data showed that approximately 70% of patients with a provisional diagnosis of lateralizing PA in our Thai cohort harbored *KCNJ5* mutations^55^, a prevalence comparable to that reported in other Asian studies^56–59^, supporting the validity of our subtype classification. Sixth, our study had heterogeneity of site-specific AVS protocols. To address this, we performed sensitivity analyses to evaluate diagnostic performance across different AVS techniques and demonstrated that AVS methods did not affect the results. A seventh limitation is that the number of patients classified as having a low probability of lateralizing PA was very small, which may limit the interpretability and generalizability of findings within this subgroup. Moreover, given that our cohort was diagnosed prior to the 2025 guideline era, relatively few patients met this definition (PAC between 277-305 pmol/L [10-11 ng/dL]). Notably, these patients exhibited a high proportion of lateralizing PAs, which likely reflects additional clinical parameters suggesting lateralizing disease rather than truly representing the low-probability phenotype as currently defined. These patients may therefore represent severe PA with aldosterone variability who underwent further workup based on clinical judgment, and may not be representative of the broader low-probability population that would be identified under systematic screening according to the 2025 guidelines. As such, conclusions drawn from this small subgroup should be interpreted with caution, and larger prospective cohorts applying the 2025 criteria from the outset will be needed to better characterize outcomes in this category. However, despite the small size of the low-probability subgroup, two-thirds of these patients had lateralizing PA, highlighting the risk of misclassification and false-negative assessments of surgically curable disease. Finally, the study included only Thai patients, which may limit generalizability; however, our findings are consistent with prior studies conducted in Canada^16,17^, France^15^, and Taiwan^18^.

In conclusion, the 2025 ES probability-based frameworks did not reliably predict PA subtype, underscoring that such models are based on low-quality evidence and are not trustworthy triage tools to predict PA subtype^26^. Our findings support the guideline assertion that patients deemed to have a high-probability for lateralizing PA are likely to benefit from AVS when surgery is desired. However, relying on aldosterone suppression testing was not useful in predicting PA subtype either in the overall cohort or in the intermediate-probability group, where it exhibited poor accuracy with high false-positive and substantial false-negative rates, consistent with several recent studies^14,16–18^. Despite the widespread desire to predict PA subtype with conventionally available tests, we found no evidence that this was reasonably applicable to individual patient cases using newly endorsed approaches by the ES guidelines in this real-world setting.

## Data Availability

The data supporting the findings of this study are available upon reasonable request from the corresponding author (W.W.P.). The data are not publicly available in accordance with the conditions of the institutional review board approvals at the participating centers, which authorized data collection for this study without provisions for public release.

## Acknowledgments

We thank the teams from Phramongkutklao Hospital, Phramongkutklao College of Medicine, and Excellence Center for Diabetes, Hormones and Metabolism, King Chulalongkorn Memorial Hospital, Thai Red Cross Society, and Faculty of Medicine, Chulalongkorn University, Bangkok, Thailand.

---

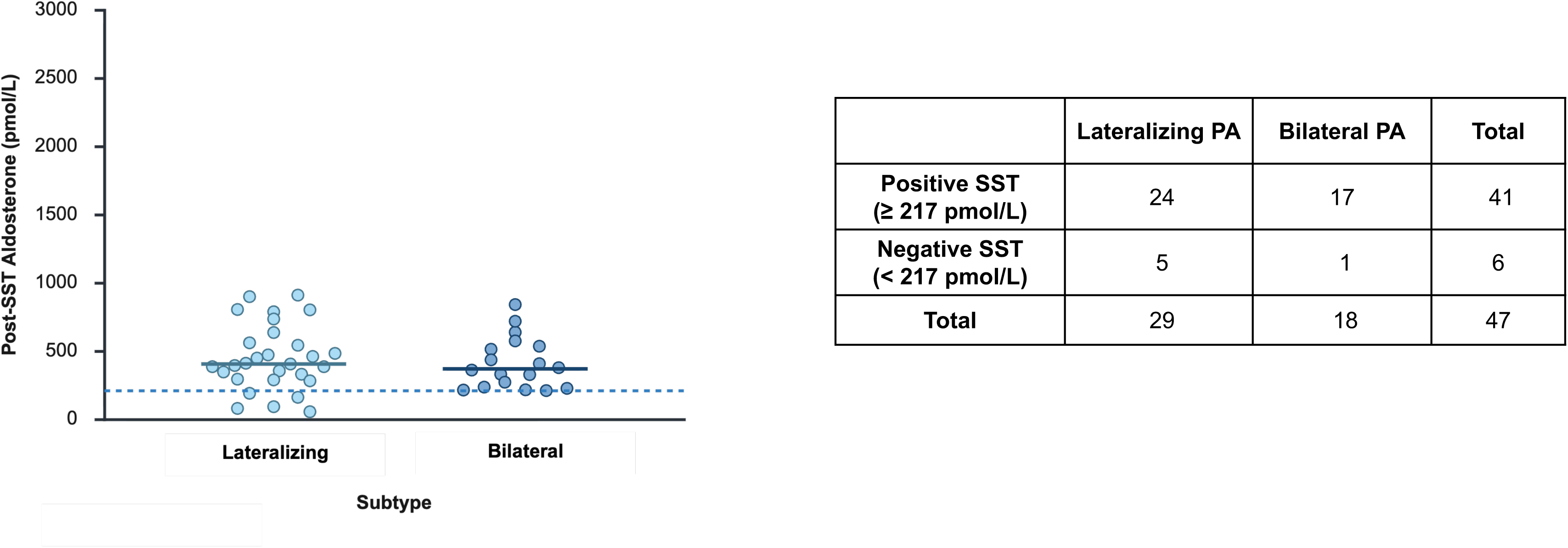

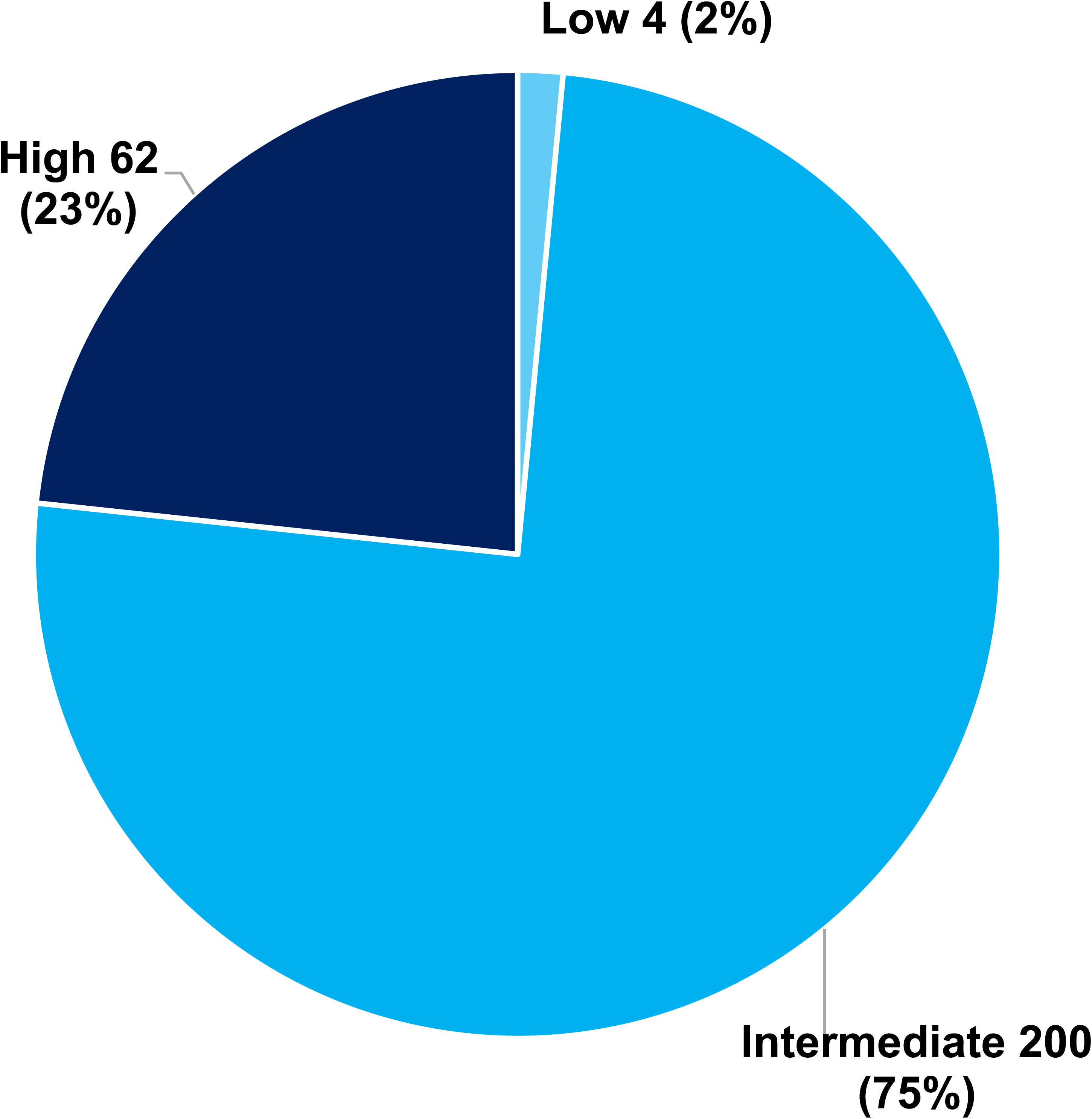

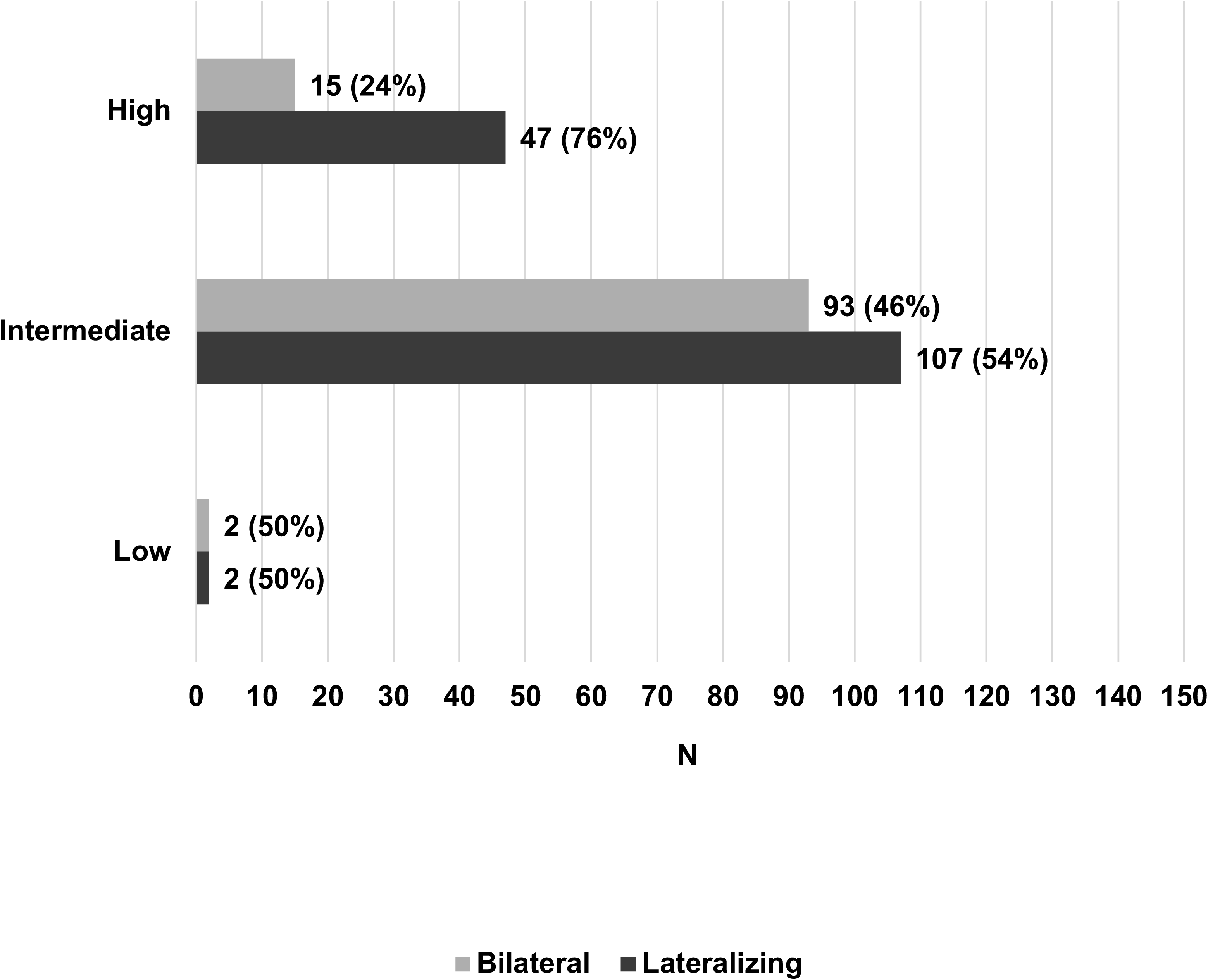

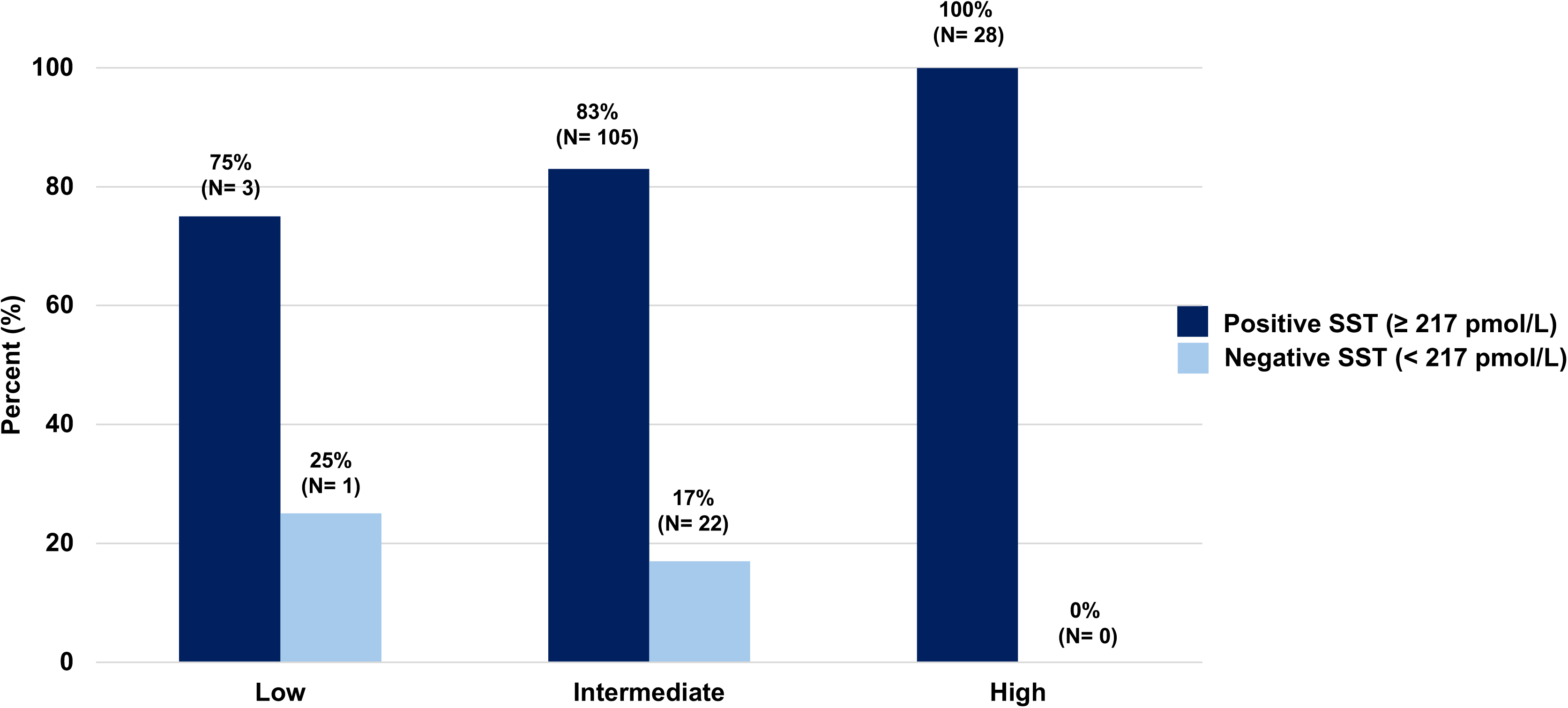

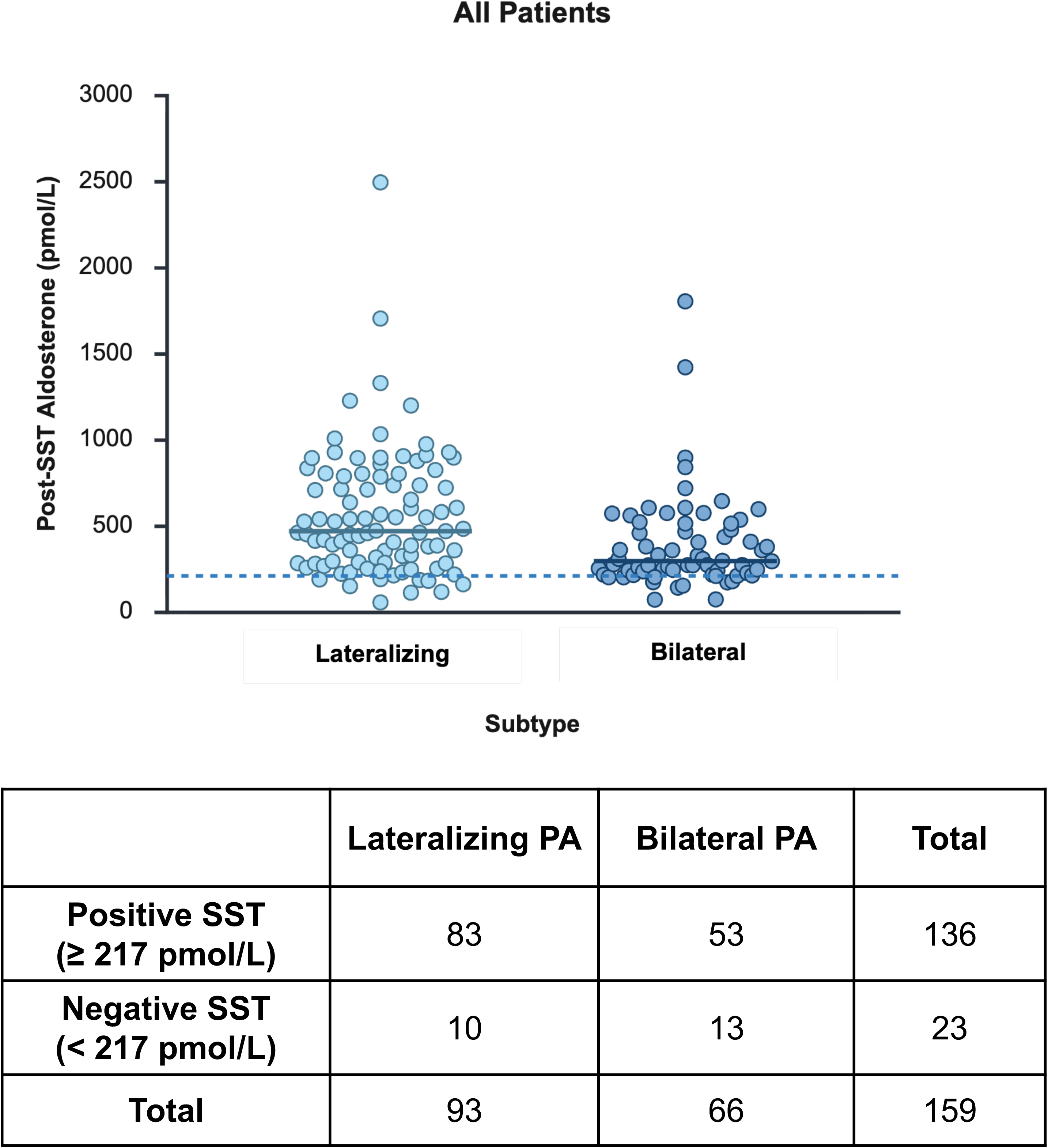

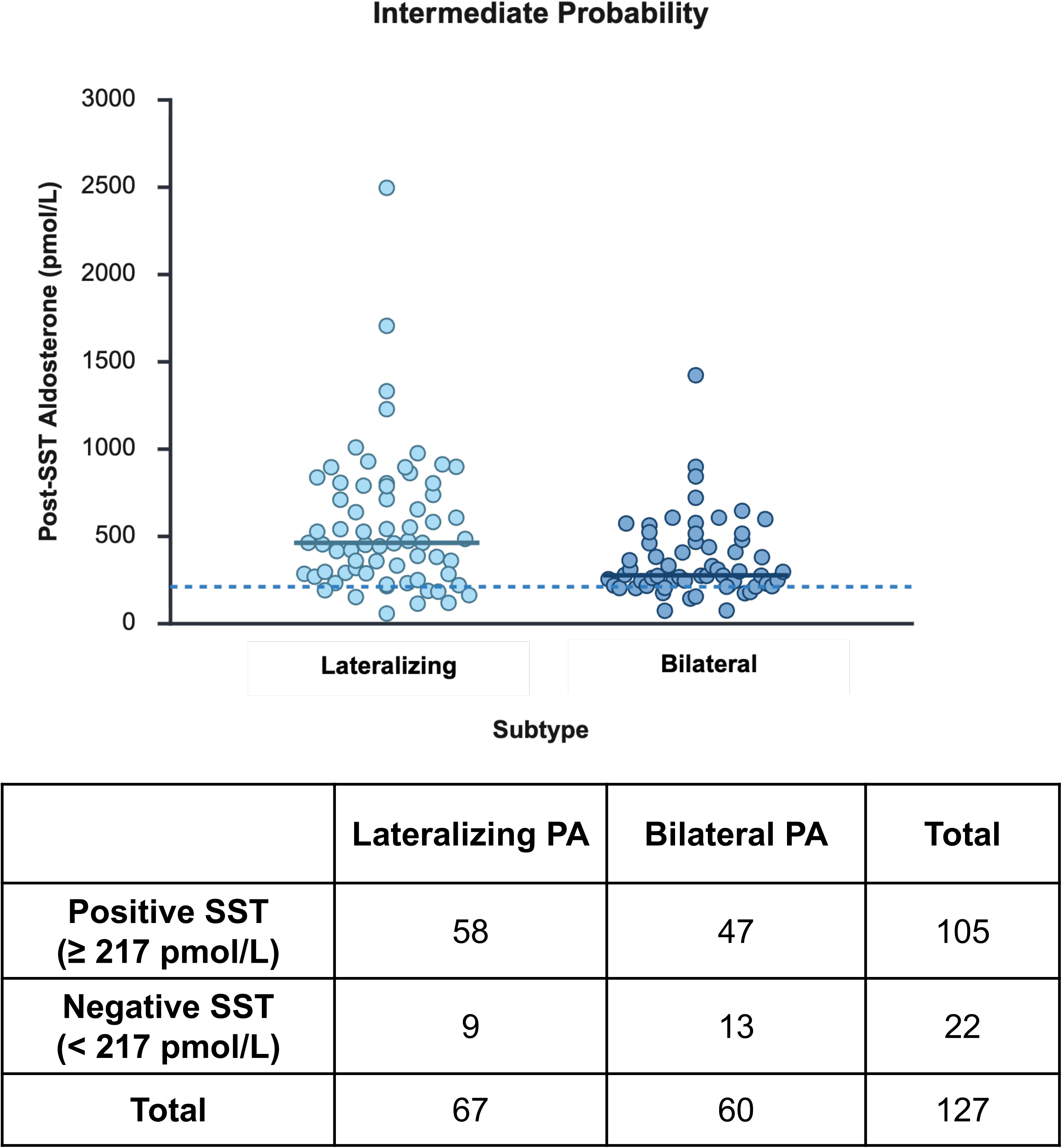

## References

1. Vaidya A, Mulatero P, Baudrand R, Adler GK. The Expanding Spectrum of Primary Aldosteronism: Implications for Diagnosis, Pathogenesis, and Treatment. Endocr Rev 2018;39(6):1057–1088. 10.1210/er.2018-00139.

2. Vaidya A, Hundemer GL, Nanba K, Parksook WW, Brown JM. Primary Aldosteronism: State-of-the-Art Review. Am J Hypertens 2022;35(12):967–988. 10.1093/ajh/hpac079.

3. Turcu AF, Yang J, Vaidya A. Primary aldosteronism - a multidimensional syndrome. Nat Rev Endocrinol 2022;18(11):665–682. 10.1038/s41574-022-00730-2.

4. Adler GK, Stowasser M, Correa RR, et al. Primary Aldosteronism: An Endocrine Society Clinical Practice Guideline. J Clin Endocrinol Metab 2025;110(9):2453–2495. 10.1210/clinem/dgaf284.

5. McEvoy JW, McCarthy CP, Bruno RM, et al. 2024 ESC Guidelines for the management of elevated blood pressure and hypertension. Eur Heart J 2024;45(38):3912–4018. 10.1093/eurheartj/ehae178.

6. Jones DW, Ferdinand KC, Taler SJ, et al. 2025 AHA/ACC/AANP/AAPA/ABC/ACCP/ACPM/AGS/AMA/ASPC/NMA/PCNA/SGIM Guideline for the Prevention, Detection, Evaluation and Management of High Blood Pressure in Adults: A Report of the American College of Cardiology/American Heart Association Joint Committee on Clinical Practice Guidelines. Hypertension 2025;82(10):e212–e316. 10.1161/hyp.0000000000000249.

7. Monticone S, Burrello J, Tizzani D, et al. Prevalence and Clinical Manifestations of Primary Aldosteronism Encountered in Primary Care Practice. J Am Coll Cardiol 2017;69(14):1811–1820. 10.1016/j.jacc.2017.01.052.

8. Parisien-La Salle S, Hundemer GL, Nehs MA, Barletta JA, Vaidya A. Treatment of Primary Aldosteronism. Hypertension 2026. 10.1161/hypertensionaha.126.26238.

9. Hundemer GL, Curhan GC, Yozamp N, Wang M, Vaidya A. Cardiometabolic outcomes and mortality in medically treated primary aldosteronism: a retrospective cohort study. Lancet Diabetes Endocrinol 2018;6(1):51–59. 10.1016/s2213-8587(17)30367-4.

10. Yang J, Burrello J, Goi J, et al. Outcomes after medical treatment for primary aldosteronism: an international consensus and analysis of treatment response in an international cohort. Lancet Diabetes Endocrinol 2025;13(2):119–133. 10.1016/s2213-8587(24)00308-5.

11. Turcu AF, Auchus R. Approach to the Patient with Primary Aldosteronism: Utility and Limitations of Adrenal Vein Sampling. J Clin Endocrinol Metab 2021;106(4):1195–1208. 10.1210/clinem/dgaa952.

12. Rossi GP, Battistel M, Seccia TM, Rossi FB, Rossitto G. Subtyping of Primary Aldosteronism by Adrenal Venous Sampling. Endocr Rev 2025;46(4):501–517. 10.1210/endrev/bnaf007.

13. Farah MH, Hegazi M, Firwana M, et al. A Systematic Review Supporting the Endocrine Society Clinical Practice Guideline on Management of Primary Aldosteronism. J Clin Endocrinol Metab 2025;110(9):e2833–e2844. 10.1210/clinem/dgaf290.

14. Parksook WW, Brown JM, Omata K, et al. The Spectrum of Dysregulated Aldosterone Production: An International Human Physiology Study. J Clin Endocrinol Metab 2024. 10.1210/clinem/dgae145.

15. Cornu E, Steichen O, Nogueira-Silva L, et al. Suppression of Aldosterone Secretion After Recumbent Saline Infusion Does Not Exclude Lateralized Primary Aldosteronism. Hypertension 2016;68(4):989–994. 10.1161/hypertensionaha.116.07214.

16. Leung AA, Padwal RS, Hundemer GL, et al. Confirmatory Testing for Primary Aldosteronism: A Study of Diagnostic Test Accuracy. Ann Intern Med 2025;178(7):948–956. 10.7326/annals-24-03153.

17. Leung AA, Padwal RS, Hundemer GL, et al. Seated Saline Suppression Test for Lateralizing Primary Aldosteronism. Hypertension 2026. 10.1161/hypertensionaha.125.26008.

18. Tsai CH, Parisien-La Salle S, Brown JM, et al. Discordance and shortcomings of aldosterone suppression tests in primary aldosteronism. Eur J Endocrinol 2025;193(3):348–358. 10.1093/ejendo/lvaf170.

19. Funder JW, Carey RM, Mantero F, et al. The Management of Primary Aldosteronism: Case Detection, Diagnosis, and Treatment: An Endocrine Society Clinical Practice Guideline. J Clin Endocrinol Metab 2016;101(5):1889–1916. 10.1210/jc.2015-4061.

20. Williams TA, Lenders JWM, Mulatero P, et al. Outcomes after adrenalectomy for unilateral primary aldosteronism: an international consensus on outcome measures and analysis of remission rates in an international cohort. Lancet Diabetes Endocrinol 2017;5(9):689–699. 10.1016/s2213-8587(17)30135-3.

21. Sukor N, Sunthornyothin S, Tran TV, et al. Health Care Challenges in the Management of Primary Aldosteronism in Southeast Asia. J Clin Endocrinol Metab 2024;109(7):1718–1725. 10.1210/clinem/dgae039.

22. Rossi GP, Crimì F, Rossitto G, et al. Feasibility of Imaging-Guided Adrenalectomy in Young Patients With Primary Aldosteronism. Hypertension 2022;79(1):187–195. 10.1161/hypertensionaha.121.18284.

23. Williams TA, Gong S, Tsurutani Y, et al. Adrenal surgery for bilateral primary aldosteronism: an international retrospective cohort study. Lancet Diabetes Endocrinol 2022;10(11):769–771. 10.1016/s2213-8587(22)00253-4.

24. Stowasser M, Eisenhofer G, Pamporaki C, Yang J, Young W, Jr. Confirmatory Testing for Primary Aldosteronism: Setting the Record Straight Under New Guidelines. Hypertension 2026;83(5):e26302. 10.1161/hypertensionaha.125.26302.

25. Leung AA, Symonds CJ, Hundemer GL, et al. Performance of Confirmatory Tests for Diagnosing Primary Aldosteronism: a Systematic Review and Meta-Analysis. Hypertension 2022;79(8):1835–1844. 10.1161/hypertensionaha.122.19377.

26. Demko J, Cohen JB. Evolving Role of Confirmatory Testing in Subtyping Primary Aldosteronism. Hypertension 2026;83(5):e26716. 10.1161/hypertensionaha.126.26716.

27. Parksook WW, Brown JM, Milks J, et al. Saline suppression testing-induced hypocalcemia and implications for clinical interpretations. Eur J Endocrinol 2024;191(2):241–250. 10.1093/ejendo/lvae099.

28. Kline GA, Leung AA, Orton D, MacFarlane J, Gurnell M. Is It Time to Retire Aldosterone Suppression Testing? Am J Hypertens 2026;39(4):473–481. 10.1093/ajh/hpaf163.

29. Lacroix A, Bourdeau I, Chasseloup F, et al. Aberrant hormone receptors regulate a wide spectrum of endocrine tumors. Lancet Diabetes Endocrinol 2024;12(11):837–855. 10.1016/s2213-8587(24)00200-6.

30. Yozamp N, Hundemer GL, Moussa M, et al. Variability of Aldosterone Measurements During Adrenal Venous Sampling for Primary Aldosteronism. Am J Hypertens 2021;34(1):34–45. 10.1093/ajh/hpaa151.

31. Kline GA, Darras P, Leung AA, So B, Chin A, Holmes DT. Surprisingly low aldosterone levels in peripheral veins following intravenous sedation during adrenal vein sampling: implications for the concept of nonsuppressibility in primary aldosteronism. J Hypertens 2019;37(3):596–602. 10.1097/hjh.0000000000001905.

32. Sam D, Kline GA, So B, et al. External Validation of Clinical Prediction Models in Unilateral Primary Aldosteronism. Am J Hypertens 2022;35(4):365–373. 10.1093/ajh/hpab195.

33. Kobayashi H, Abe M, Soma M, et al. Development and validation of subtype prediction scores for the workup of primary aldosteronism. J Hypertens 2018;36(11):2269–2276. 10.1097/hjh.0000000000001855.

34. Burrello J, Burrello A, Pieroni J, et al. Development and Validation of Prediction Models for Subtype Diagnosis of Patients With Primary Aldosteronism. J Clin Endocrinol Metab 2020;105(10). 10.1210/clinem/dgaa379.

35. Ng E, Gwini SM, Zheng W, Fuller PJ, Yang J. Tools to Predict Unilateral Primary Aldosteronism and Optimise Patient Selection for Adrenal Vein Sampling: A Systematic Review. Clin Endocrinol (Oxf) 2025;103(1):3–12. 10.1111/cen.15225.

36. Nanba K, Tsuiki M, Umakoshi H, et al. Shortened saline infusion test for subtype prediction in primary aldosteronism. Endocrine 2015;50(3):802–806. 10.1007/s12020-015-0615-9.

37. Kaneko H, Umakoshi H, Ishihara Y, et al. Seated saline infusion test in predicting subtype diagnosis of primary aldosteronism. Clin Endocrinol (Oxf) 2019;91(6):737–742. 10.1111/cen.14111.

38. Kološová B, Waldauf P, Wichterle D, et al. Validation of Existing Clinical Prediction Tools for Primary Aldosteronism Subtyping. Diagnostics (Basel) 2022;12(11). 10.3390/diagnostics12112806.

39. Murakami M, Naruse M, Kobayashi H, et al. Expert Consensus on the Primary Aldosteronism Severity Classification and its strategic application in indicating adrenal venous sampling. Eur J Endocrinol 2025;193(1):85–96. 10.1093/ejendo/lvaf117.

40. Eisenhofer G, Durán C, Cannistraci CV, et al. Use of Steroid Profiling Combined With Machine Learning for Identification and Subtype Classification in Primary Aldosteronism. JAMA Netw Open 2020;3(9):e2016209. 10.1001/jamanetworkopen.2020.16209.

41. Williams TA, Constantinescu G, Pamporaki C. Biomarkers for subtype stratification in primary aldosteronism: current and future perspectives. Eur J Endocrinol 2025;193(5):R51–r64. 10.1093/ejendo/lvaf218.

42. Tsai CH, Kong PH, Hsieh CC, et al. Proteomic signatures to detect unilateral primary aldosteronism in hypertensive patients. Eur J Clin Invest 2025;55(11):e70081. 10.1111/eci.70081.

43. Teo AED, Tran HTN, Khoo CM, et al. Approach to the Patient With Primary Aldosteronism: Role of Molecular Imaging. J Clin Endocrinol Metab 2025;110(12):3559–3568. 10.1210/clinem/dgaf396.

44. Årstad E, Sander K, Kurzawinski TR, et al. Adrenal Aldosterone Synthase Expression Imaging in Primary Aldosteronism. N Engl J Med 2025;393(21):2168–2170. 10.1056/NEJMc2507481.

45. Vaidya A. The Promise of Nuclear Imaging as an Alternative to Adrenal Venous Sampling for the Detection of Aldosterone-Producing Adenomas. J Clin Endocrinol Metab 2023. 10.1210/clinem/dgad542.

46. Long T, Liu G, Zhou M, et al. First-in-Human Evaluation of [ 18 F]AldoView: A Highly Selective PET Tracer for Aldosterone Synthase Imaging in Primary Aldosteronism. Clin Nucl Med 2025;50(9):847–855. 10.1097/rlu.0000000000006014.

47. Cohen JB. Rethinking Confirmatory Testing in Primary Aldosteronism. Ann Intern Med 2025;178(7):1044–1045. 10.7326/annals-25-01368.

48. Yozamp N, Hundemer GL, Moussa M, et al. Intraindividual Variability of Aldosterone Concentrations in Primary Aldosteronism: Implications for Case Detection. Hypertension 2021;77(3):891–899. 10.1161/hypertensionaha.120.16429.

49. Ng E, Gwini SM, Libianto R, et al. Aldosterone, Renin, and Aldosterone-to-Renin Ratio Variability in Screening for Primary Aldosteronism. J Clin Endocrinol Metab 2022;108(1):33–41. 10.1210/clinem/dgac568.

50. Maciel AAW, Freitas TC, Fagundes GFC, et al. Intra-individual Variability of Serum Aldosterone and Implications for Primary Aldosteronism Screening. J Clin Endocrinol Metab 2023;108(5):1143–1153. 10.1210/clinem/dgac679.

51. Ng E, Gwini SM, Stowasser M, et al. Aldosterone and renin concentrations and blood pressure in young Indigenous and non-Indigenous adults in the Northern Territory: a cross-sectional study. Med J Aust 2023;219(6):263–269. 10.5694/mja2.52062.

52. Hundemer GL, Leung AA, Kline GA, Brown JM, Turcu AF, Vaidya A. Biomarkers to Guide Medical Therapy in Primary Aldosteronism. Endocr Rev 2024;45(1):69–94. 10.1210/endrev/bnad024.

53. Grundy MC, Leung AA, Pasieka JL, et al. Outcomes After Unilateral Adrenalectomy in Asymmetrical Bilateral Primary Aldosteronism. Hypertension 2025;82(10):1612–1622. 10.1161/hypertensionaha.125.24849.

54. Sukor N, Gordon RD, Ku YK, Jones M, Stowasser M. Role of unilateral adrenalectomy in bilateral primary aldosteronism: a 22-year single center experience. J Clin Endocrinol Metab 2009;94(7):2437–2445. 10.1210/jc.2008-2803.

55. Warachit W, Atikankul T, Houngngam N, Sunthornyothin S. Prevalence of Somatic KCNJ5 Mutations in Thai Patients With Aldosterone-Producing Adrenal Adenomas. J Endocr Soc 2018;2(10):1137–1146. 10.1210/js.2018-00097.

56. Zheng FF, Zhu LM, Nie AF, et al. Clinical characteristics of somatic mutations in Chinese patients with aldosterone-producing adenoma. Hypertension 2015;65(3):622–628. 10.1161/hypertensionaha.114.03346.

57. Wu VC, Huang KH, Peng KY, et al. Prevalence and clinical correlates of somatic mutation in aldosterone producing adenoma-Taiwanese population. Sci Rep 2015;5:11396. 10.1038/srep11396.

58. Okamura T, Nakajima Y, Katano-Toki A, et al. Characteristics of Japanese aldosterone-producing adenomas with KCNJ5 mutations. Endocr J 2017;64(1):39–47. 10.1507/endocrj.EJ16-0243.

59. Nanba K, Rainey WE. GENETICS IN ENDOCRINOLOGY: Impact of race and sex on genetic causes of aldosterone-producing adenomas. Eur J Endocrinol 2021;185(1):R1–r11. 10.1530/eje-21-0031.

